# Resting-state background features demonstrate multidien cycles in long-term EEG device recordings

**DOI:** 10.1101/2023.07.05.23291521

**Authors:** William K.S. Ojemann, Brittany H. Scheid, Sofia Mouchtaris, Alfredo Lucas, Joshua J. LaRocque, Carlos Aguila, Arian Ashourvan, Lorenzo Caciagli, Kathryn A. Davis, Erin C. Conrad, Brian Litt

## Abstract

**Background:** Longitudinal EEG recorded by implanted devices is critical for understanding and managing epilepsy. Recent research reports patient-specific, multi-day cycles in device-detected epileptiform events that coincide with increased likelihood of clinical seizures. Understanding these cycles could elucidate mechanisms generating seizures and advance drug and neurostimulation therapies

**Objective/Hypothesis:** We hypothesize that seizure-correlated cycles are present in background neural activity, independent of interictal epileptiform spikes, and that neurostimulation may disrupt these cycles.

**Methods:** We analyzed regularly-recorded seizure-free data epochs from 20 patients implanted with a responsive neurostimulation (RNS) device for at least 1.5 years, to explore the relationship between cycles in device-detected interictal epileptiform activity (dIEA), clinician-validated interictal spikes, background EEG features, and neurostimulation.

**Results:** Background EEG features tracked the cycle phase of dIEA in all patients (AUC: 0.63 [0.56 - 0.67]) with a greater effect size compared to clinically annotated spike rate alone (AUC: 0.55 [0.53-0.61], p < 0.01). After accounting for circadian variation and spike rate, we observed significant population trends in elevated theta and beta band power and theta and alpha connectivity features at the cycle peaks (sign test, p < 0.05). In the period directly after stimulation we observe a decreased association between cycle phase and EEG features compared to background recordings (AUC: 0.58 [0.55-0.64]).

**Conclusions:** Our findings suggest that seizure-correlated dIEA cycles are not solely due to epileptiform discharges but are associated with background measures of brain state; and that neurostimulation may disrupt these cycles. These results may help elucidate mechanisms underlying seizure generation, provide new biomarkers for seizure risk, and facilitate monitoring, treating, and managing epilepsy with implantable devices.

**Highlights:**

- Background EEG features track multidien cycles in RNS dIEA
- dIEA linked EEG features are patient-specific and may differ with cortical structure
- Responsive neurostimulation suppresses EEG-dIEA coupling

## Introduction

Temporal cycles in seizures have been documented extensively in animals and humans for many years [1]–[4]. Recently, recordings from chronic implantable EEG recording devices have yielded new insights into seizure generation in patients with epilepsy [5], [6]. In particular, Baud et al. found patient-specific, periodic multi-day (“multidien”) fluctuations in device-detected interictal epileptiform activity (dIEA) over the course of weeks to months in long-term recordings from the implantable responsive neurostimulation (RNS) device [5]. More recently, similar cycles have been described in long-term EEG recordings from other devices in humans [6]–[8], dogs [9], and rats [10], and in longitudinal measures of heart rate [11].

Device-detected IEA counts are based on combinations of detection parameters that are hand-tuned by neurologists to be sensitive to acute aberrant brain activity. In practice, these detection parameters vary over time, and it is unclear how often they track epileptiform discharges or some other phenomena. Despite the heterogeneous and subjective nature of these detection settings, there is evidence that tracking patient-specific dIEA can be useful as a predictive biomarker of seizure risk [5], [12]. While numerous theories exist regarding the origin of multidien cycles, and early evidence indicates that cycles can be perturbed with neuromodulation [7], little is known about how their biological underpinnings relate to neural function and therapy [4].

Understanding these cycles and their relationship to other measures of brain state would allow us to discover novel ways to titrate treatment and support patient-specific, phase-locked therapeutic paradigms. Prior efforts to investigate multidien fluctuations in long-term neural recordings are based upon measures of acute epileptiform activity (i.e. spikes, dIEA) [5], [8], [13], however it is unclear whether these cycles result from epileptiform activity or some other phenomena. The relationship between dIEA cycles and background EEG measures remains unexplored.

In this study we investigate the relationship between interictal background EEG features and dIEA cycle phase. We test the hypothesis that the dIEA cycle that best aligns with periodic seizure occurrence is represented in background neural activity as well as in clinical spike rate, and that neurostimulation suppresses this relationship. To test these hypotheses, we measure associations between dIEA cycle phase and features calculated from interictal recordings: spike rate, band power, and connectivity both at baseline and in a window following stimulation. We identify novel associations between seizure-associated dIEA cycles and measures of background brain state, suggesting that the observed cycles are not solely based on acute discharges and creating a platform for generating novel hypotheses that further our understanding and treatment of epilepsy.

## Materials and Methods

### RNS system recordings

The RNS System is a neuromodulation therapy that uses an implantable device with two bipolar channels each in two electrode leads (4 channels total). EEG activity in two selected detection channels is continuously monitored, and the device responds with electrical stimulation pulses when abnormal activity is detected [14]. A count of hourly detections is recorded by the device and has previously been referred to as a measure of interictal epileptiform activity (IEA) [5]. Here we denote device-detected interictal epileptiform activity as “dIEA”, to distinguish it from clinically-verified epileptiform spikes that we extract from EEG recordings (in methods below).

In addition to recording counts of hourly dIEA detections, the RNS device records 90-second “Scheduled Event” (SE) EEG recordings every 12 hours to capture baseline activity (**Figure 1D**), and 90-second “Long Episode” (LE) recordings, which capture likely seizures. Due to device memory limitations, SE clips are often set as low priority and can be overwritten by other stored EEG clips. Because the detection criteria are met so frequently, SEs often capture stimulations as well as baseline activity (**Figure 1F**). In this study, we used hourly dIEA detections, LE capture timepoints, and EEG signals recorded in SE events both with and without stimulation.

**Figure 1.**
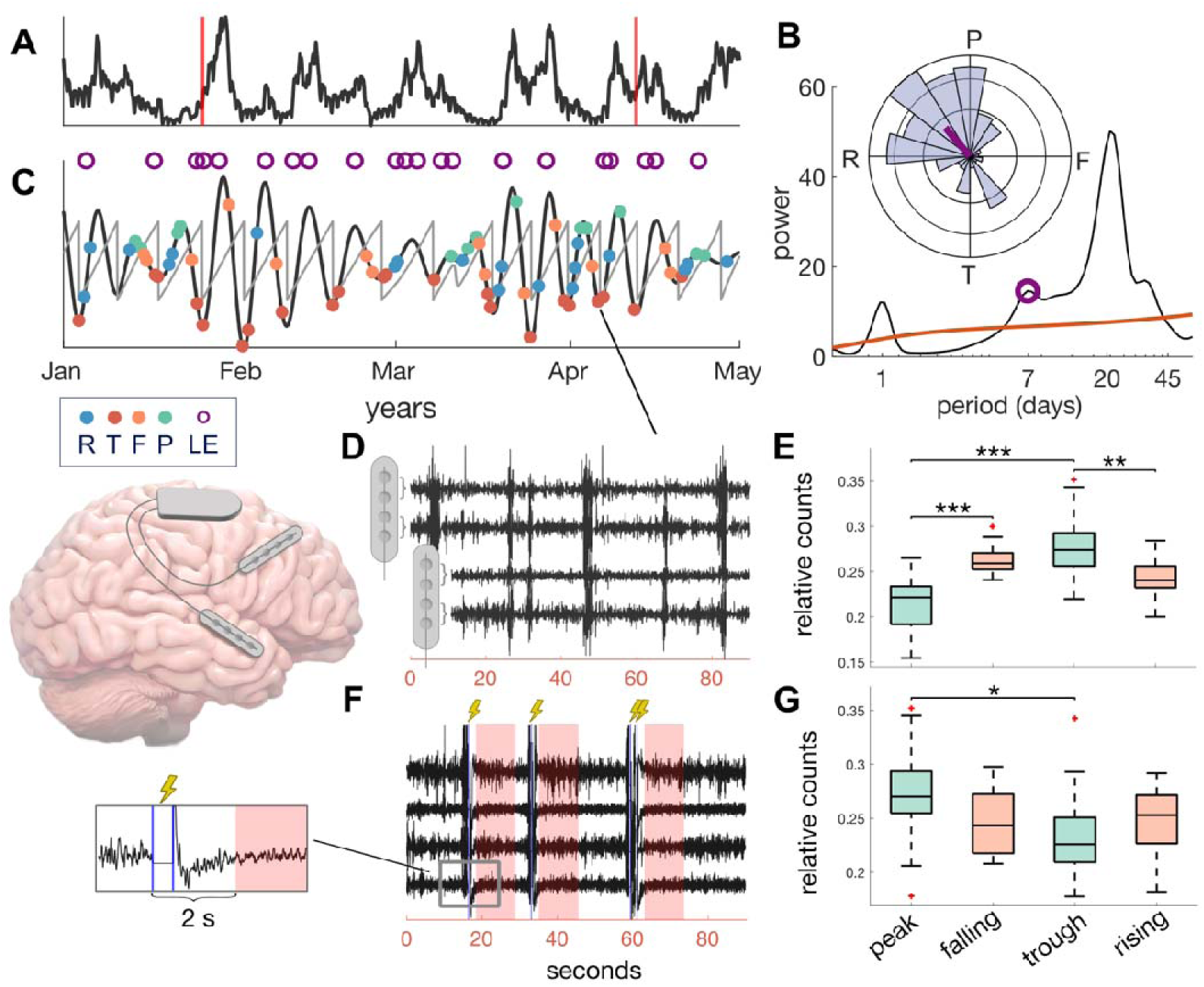
EEG clip selection and dIEA phase labeling. We illustrate our methods in panels A-E using data from the same representative patient (HUP096). **A)** Hourly counts of detected interictal epileptiform activity (dIEA) after z-scoring between patient visits (vertical red lines). A 100-h moving average was applied for visualization. **B)** The periodogram obtained by averaging the spectrogram of the dIEA signal over time. The peak associated with the greatest LE PLV is indicated by the purple marker. **Inset:** Histogram showing mean multidien phase and axial mean of LE occurrences. LEs in this patient were phase-locked to the rising phase of the multidien cycle. **C)** Signal component after wavelet decomposition of dIEA signal with period corresponding to the marked periodogram peak in **B**. Sawtooth lines indicate instantaneous phase, purple markers indicate LE recordings, filled dots indicate sampling of SE recordings in peak (P), falling (F), trough (T), and rising (R) phase bins. **D)** Example SE recording without stimulation sampled at the cycle trough. Each channel represents a bipolar montage between pairs of adjacent contacts on the same electrode. **E)** Distribution of recorded stimulation-free SEs across each phase bin. **F)** Example SE recording with shaded 10-s analysis windows following simulations marked in blue. **G)** Distribution of post-stimulation analysis windows across each phase bin. LE-long episode, PLV-phase-locking value, SE-scheduled event, R-rising, P-peak, F-falling, T-trough

### Patient population

We retrospectively analyzed longitudinal RNS EEG and dIEA recordings from twenty out of 28 patients with drug-resistant epilepsy who were implanted with the RNS System (NeuroPace, Inc., Mountain View, CA) at the Hospital at the University of Pennsylvania. We included patients if they had at least one year of dIEA recordings (recordings could be segmented as long as segments were at least three months in duration). Patients were required to have at least 200 recorded SE clips without stimulation, or 200 post-stimulation analysis windows, so that enough samples were present for our multivariate analysis. We also excluded patients without a significant multidien cycle in their dIEA counts. Eight patients did not meet our inclusion criteria (full breakdown is detailed in the **Supplemental Methods**). The remaining 20 patients included in this study were implanted with the RNS device between August 2015 and January 2022. Patient demographics are summarized in **Table 1**. All patients considered for this study gave written informed consent in accordance with the Institutional Review Board of the University of Pennsylvania.

**Table 1.** Patient Demographics.

| ID | sex | age at onset | baseline freq (sz/wk) | lead laterality | implant region | age at implant | lead locations | duration analyzed (yrs) | % interp. | % discarded | LE PLV | cycle period |
| --- | --- | --- | --- | --- | --- | --- | --- | --- | --- | --- | --- | --- |
| HUP047 | M | 10-19 | 87.5 | R | N | 50-59 | frontal | 4.38 | 0 | 2.05 | 0.16 | 31.17 |
| HUP084 | M | < 10 | 2.57 | B | M | 50-59 | L. Hipp/ R. Hipp | 4.92 | 0 | 1.65 | 0.17 | 12.37 |
| HUP096 | F | 30-39 | 6.42 | B | N | 50-59 | temporal | 2.84 | 0.016 | 2.82 | 0.40 | 6.94 |
| HUP108† | M | 10-19 | 1.75 | B | M | 30-39 | L. Hipp/ R. Hipp | 3.36 | 0 | 2.40 | 0.24 | 7.35 |
| HUP109 | M | 40-49 | 70 | B | M | 60-69 | L. Hipp/ R. Hipp | 2.15 | 0 | 3.69 | 0.18 | 31.17 |
| HUP127 | F | 10-19 | 2.5 | B | M | 30-39 | L. Hipp/ R. Hipp | 3.52 | 0 | 2.29 | 0.17 | 41.6 |
| HUP128 | F | 10-19 | 8.5 | L | N | 60-69 | temporal | 2.32 | 0 | 9.30 | 0.05 | 49.47 |
| HUP129 | M | 30-39 | 1.5 | R | B | 40-49 | R. Hipp/ R Insula | 3.09 | 0 | 2.60 | 0.07 | 24.74 |
| HUP131 | M | <10 | 17.5 | L | B | 20-29 | frontal | 3.41 | 0 | 4.28 | 0.24 | 46.7 |
| HUP136 | F | 40-49 | 0.11 | L | B | 50-59 | L. Frontal/ L Temporal | 5.54 | 0 | 1.47 | 0.38 | 13.88 |
| HUP137 | M | 30-39 | 3.15 | B | M | 50-59 | L. Hipp/ R. Hipp | 3.72 | 0 | 2.17 | 0.42 | 11.02 |
| HUP143* | F | 10-29 | 1.05 | B | M | 20-29 | L. Hipp/ R. Hipp | 3.25 | 0 | 2.47 | 0.36 | 10.4 |
| HUP147* | F | 10-29 | 14 | L | N | 40-49 | parietal/insula | 3.35 | 0 | 2.40 | 0.18 | 44.08 |
| HUP153 | F | 40-49 | 0.58 | B | M | 50-59 | L. Hipp/ R. Hipp | 3.12 | 0 | 2.57 | 0.27 | 29.42 |
| HUP156 | F | 10-19 | 0.23 | L | N | 40-49 | temporal | 2.76 | 0 | 2.90 | 0.49 | 13.88 |
| HUP197* | F | < 10 | 2 | L | N | 40-49 | Temporal | 2.43 | 0 | 3.28 | 0.04 | 23.35 |
| RNS021 | F | < 10 | 0.58 | B | M | 10-19 | L. Hipp/ R. Hipp | 1.63 | 0 | 4.82 | 0.24 | 26.21 |
| RNS022 | F | 10-19 | 1.87 | B | M | 30-39 | L. Hipp/ R. Hipp | 3.36 | 0 | 5.82 | 0.34 | 16.51 |
| RNS026 | M | 20-29 | 0.23 | B | M | 20-29 | L. Hipp/ R. Hipp | 2.37 | 0 | 3.36 | 0.18 | 17.49 |
| RNS029 | F | 20-29 | 0.58 | B | M | 40-49 | L. Hipp/ R. Hipp | 4.58 | 0 | 1.77 | 0.27 | 20.8 |
| **mean (std)** |  | 20.4 (13.5) | 11.1 (23.8) |  |  | 43.8 (12.7) |  | 3.3 (0.9) |  | 3.2 (1.8) | 0.24 (0.13) | 23.9 (13.3) |

\*non-stimulation analysis only, †post-stimulation analysis only, L/R/B: left/right/bilateral, M/N/B: mesial temporal, neocortical, both, % interp: percent of interpolated dIEA signal, % discarded: percent of dIEA recording omitted due to implant effect or discontinuity.

### Extracting multidien cycles from dIEA counts

We followed a similar methodology as outlined in Baud et al. to extract multidien cycles from dIEA activity (**Figure 1A**) [5], [15]. In brief, for each patient we applied a wavelet transform to the dIEA signal and identified the dominant modes in the average frequency spectrum (peaks in the periodogram) in the range of 3-60 days. We performed a wavelet decomposition at each peak periodicity and for each patient, we used the wavelet component that was most phase-locked to seizure occurrence for our analysis (**Figure 1B, C**). Finally, we calculated phase angle by applying the Hilbert transform to each patient’s extracted multidien cycle (**Figure 1C**). See **Supplemental Methods** for more details.

### Extracting electrophysiology from RNS EEG recordings

#### Selecting device EEG recordings

We analyzed only SE device recordings because we were interested in understanding the relationship between *baseline* brain state and dIEA cycle phase. We divided our analysis of SEs into two separate datasets: (1) SEs without detections and subsequent simulations (N=19 patients), and (2) SEs containing simulations followed by a 12-second window of stimulation-free EEG (N = 17 patients) (**Figure 1D, F**). SEs are non-uniformly distributed in time (**Figure S13**) precluding us from applying circular analyses and wavelet decompositions to directly compare EEG recordings with dIEA cycles. We assigned a phase angle to each SE based on its phase location within the extracted dominant multidien cycle. The SE clips were categorized into four phase bins: peak (-π/4 to π/4), falling (π/4 to 3π/4), trough (3π/4 to -3π/4), and rising (-3π/4 to -π/4).

#### Extracting spike rate

We detected spikes in all SE clips using a modified version of a previously validated spike detector [16] (**Figure 2A**) (see **Supplemental Methods** for detector parameters). One board-certified neurologist (JL) independently validated a random selection of 50 spikes per patient. The median positive predictive value (PPV) across patients was 70% (IQR: [47.5%, 78.5%]).

**Figure 2.**
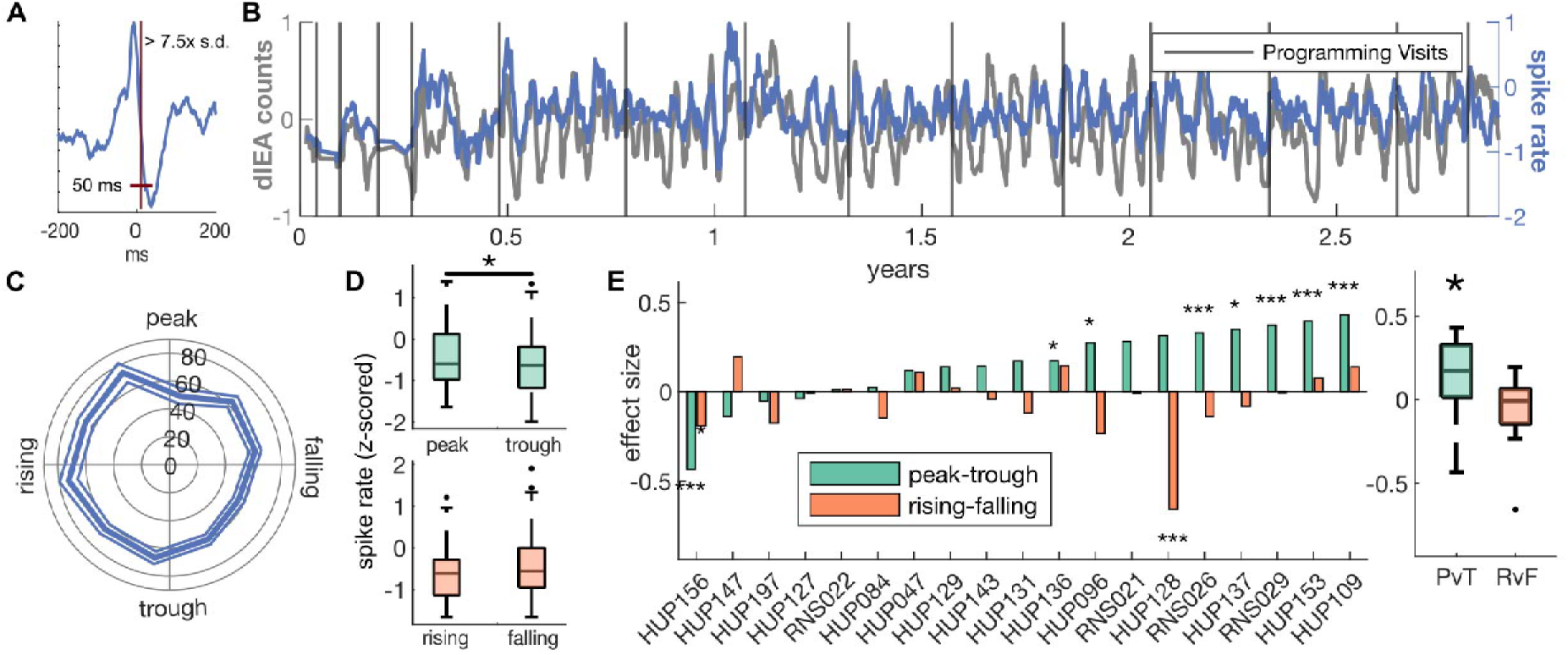
Spike rate association with dIEA counts and multidien cycles. A) Illustration of spike detection limits for an example spike detection. B) Spike rate in scheduled events and dIEA counts for sample subject HUP096. For visualization purposes, dIEA counts were smoothed using a moving average with window size equal to the median schedule event sampling rate (∼12 hours) and the averaged dIEA signal was resampled to correspond with the recorded scheduled events. C) Polar plot for sample subject HUP096 of average spike rate (spikes/90s clip) and standard error across phases of their multidien cycle. D) Box plots show distribution of spike rate in rising/falling (green) and peak/trough (orange). E) Cohen’s D effect sizes of spike rate association in PvT (green) or RvF (orange). Star represent a significant difference in phase groups (signtest). Population distribution of effect sizes i represented in the box plot at right, with the distribution of PvT effect sizes being significantly greater than zero (signtest).

#### Extracting representative brain-state features

We calculated band power and connectivity in four frequency bands - theta (4-8 Hz), alpha (8-13 Hz), beta (13-30 Hz), and gamma (33-100 Hz) (**Table 2**). In SEs without stimulation, we calculated features across the entire 90s clip, in SEs with stimulation, features were calculated within the 10-second window beginning two seconds after the stimulation occurred (**Figure 1F)** [17]. We calculated the maximum feature value between detection channels (band power) and between leads (connectivity) as a summary statistic for each feature in each window. We also calculated a binary time-of-day feature, delimited at 8am/8pm, to account for circadian variation in the features when analyzing their model coefficients. To address changes in detection parameters and baseline drifts in the features, we z-scored each feature between clinical neurologist visits. See the **Supplementary Methods** for details on feature calculations.

**Table 2:**
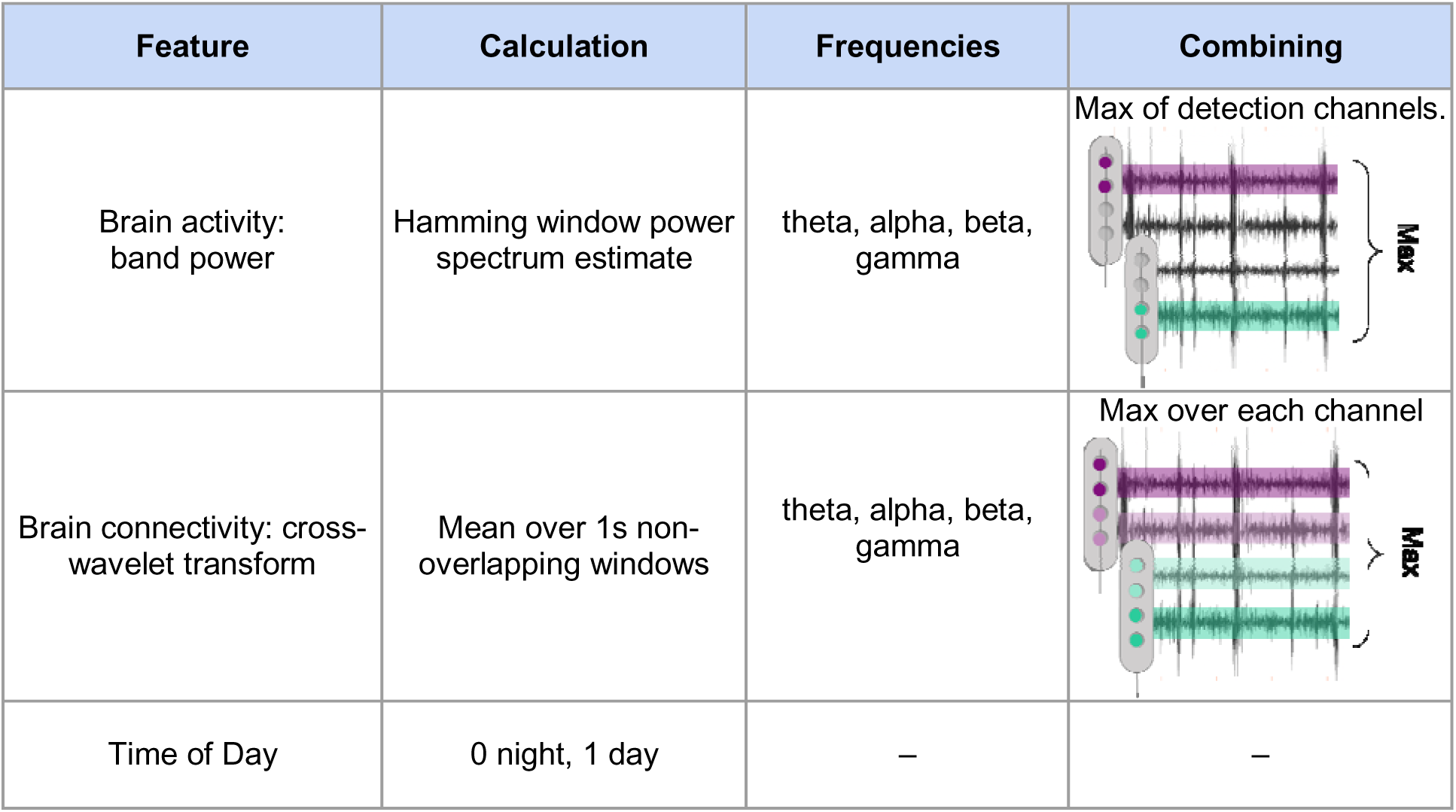
Feature calculations. Two types of features were extracted from the EEG recordings measuring (1) neural activity at each detection channel and (2) functional connectivity within leads. Each feature was calculated in four canonical frequency bands encapsulating the entire frequency spectrum captured by the RNS device. For each bandpower feature we took the maximum value between the two detection channels (purple vs green). For each connectivity feature we took the maximum value between the two leads (purple vs green). We also included time of day of the recording to account for potential circadian variations when interpreting our features.

| Feature | Calculation | Frequencies | Combining |
| --- | --- | --- | --- |
| Brain activity: band power | Hamming window power spectrum estimate | theta, alpha, beta, gamma | Max of detection channels. |
| Brain connectivity: cross-wavelet transform | Mean over 1s non-overlapping windows | theta, alpha, beta, gamma | Max over each channel |
| Time of Day | 0 night, 1 day | — | — |

### Statistical analysis

#### Multidien cycle significance and phase distribution

We tested the significance of the dIEA periodogram peak associated with the highest LE PLV value using the 99% confidence interval of an autoregression-based red noise null model for periodic signals, a method previously applied to find multidien cycles in humans [18], [19]. When analyzing the phase entrainment of LEs to cycle phase, we used an omnibus test for non-uniformity to assess the significance of LE phase locking [5], [20].

#### Univariate feature analysis

For each univariate feature extracted from the SEs, we used a one-way ANOVA to test for significant differences in feature values across the four phase bins. We further split the feature comparison into two groups to increase interpretability: peak vs. trough (PvT) phase bins, and rising vs. falling (RvF) phase bins. We calculated the effect size (Cohen’s *d*) between the peak and trough groups, and the rising and falling groups. We report effect size as Cohen’s *d* to better characterize both the directionality and sample-size-independent magnitude of the relationships[21]. We tested for group-level trends in effect-size and directionality across the patient population using a two-tailed sign test against a zero median for each feature (**Figure S5**).

#### Multivariate machine learning model

We developed a multivariate classification model to understand how band power and connectivity features relate to dIEA cycle phase. For each patient, we built two linear support vector machines (SVMs) which used the EEG features to classify PvT and RvF phases, respectively. We transformed the features using principal component analysis (PCA) to account for multicollinearity. For each patient-specific PvT and RvF model, we performed 10-fold cross validation to obtain a robust estimate of model performance. We report the mean area under the receiver operating curve (AUROC) across all of the k-folds as a measure of generalization performance effect size. To assess the statistical significance of the AUROC values, we used the p-value of the associated Mann-Whitney U statistic comparing the predicted scores between the true negative (trough/falling) and true positive (peak/rising) samples (**Figure S5,8**) [22].

We analyzed the magnitude and direction of the model coefficients to better understand the relationship between the EEG features and cycle phase. We applied the inverse PCA transform to the model coefficients to obtain individual feature importances. We tested for group-level trends in coefficient magnitude across the patient population using a two-tailed sign test against a zero median for each feature.

#### General Statistical Tests

For all paired (unpaired) non-parametric tests we used a sign test of differences (rank sum) to compare model effect sizes between patients. We used spearman correlation to analyze population trends between periodogram attributes and model performance. When comparing distributions within patients we applied parametric statistical tests for both paired and unpaired analyses. We tested for an uneven distribution of SEs at the population level both with and without stimulations across the four phase bins using a Kruskal-Wallis test, and for each individual we used a chi-square goodness of fit test against a uniform distribution to test for class imbalance. For all box plots, the middle line indicates the median, box edges are lower and upper quartiles, and whisker edges are the maximum and minimum. For all assessments, the significance threshold (α) was set at 0.05, and significance is indicated in figures with *= p < 0.05, **=p < 0.01, and *** = p < 0.001. Because this paper is largely exploratory, we do not correct for multiple comparisons.

## Results

### Multidien cycles in interictal epileptiform activity

In our retrospective cohort of 20 patients, we replicated analyses from previous studies [3], [5], [15] to compare the multidien dIEA cycle characteristics in our subjects with existing literature. We found significant multidien cycles in 89% of our patient population, with patient-specific cycle periods ranging from 7 to 49 days, consistent with prior studies [4], [5]. On average, subjects possessed 2 ± 0.65 significant peaks with a median significant multidien cycle length of 22.08 [13.13, 31.17] (**Figure S1**). Our analysis of the LE phase-locked cycles showed that the distribution of seizures across cycle phase was significantly non-uniform in 18 of 20 patients, with a median PLV of 0.24 [.17, .35], consistent with previously reported values (**Figure S2**)[4], [5]. We observed a significant negative correlation (R: -0.603, p = 0.005) between the peak period length of the LE phase-locked cycle and the LE phase-locking value, indicating stronger phase-locking in shorter multidien cycles (**Figure S3**).

Because dIEA cycles represent fluctuations in stimulation event frequency, we quantified the disparity in the distribution of SE events both with *and* without stimulation across the four phase bins, given the higher number of stimulations at the cycle peak. Group-level analysis showed a significant difference in the distribution of available SE clips without stimulation across phase groups, with the largest difference between peak and trough recordings (KW test (72); Chi-sq = 33.8, p = 2.18e-07) (**Figure 1E**). When analyzing SEs containing stimulations, we found a smaller group-level class imbalance in the opposite direction (more peak than trough recordings) (KW test (67) Chi-sq = 8.53, p = 0.0363) (**Figure 1G**), (**Figure S4**). These results demonstrate distinct distributions between baseline SE recordings with and without stimulation.

### Spike rate is associated with dIEA counts

We found that patients demonstrated a moderate correlation (Pearson *r* > 0.3) between spike rate and dIEA counts with a medium effect size (median *r*: 0.34, IQR: [.23, .48]). We next asked to what extent the spike rate extracted from the device recordings could differentiate between cycle phases. At the population level, we found that spike rate was greater at cycle peaks vs. troughs (sign test U(19)=15, p=0.0192), with 72% (15/19) of patients showing a higher spike rate at cycle peaks. Univariate effect sizes were not correlated with the positive predictive value of the spike detector (p > 0.5). We did not observe a significant difference at the population level in spike rate between the rising and falling dIEA cycle phases (**Figure 2E**). The spike rates we analyzed were calculated outside of SE recordings containing detected events, suggesting that the dIEA cycle is in part measuring the brain state that is associated with epileptiform spike rate in background recordings.

### Phase-dependence of interictal EEG features

We asked whether features of background interictal activity were related to dIEA cycle phase (**Figure S5**). Our one-way ANOVA revealed that individual patients had at least one significant feature that was strongly associated with phase, though not the same feature for all patients. The proportion of day versus night recordings was similar across phase groups. The full results of the patient-specific ANOVA models are described in **Table S1 & Figure S7**.

We next split our analysis into two paired comparisons, peak vs. trough and rising vs. falling, because we expected to see the largest differences in features between the peaks and the troughs of the cycles. For the PvT comparison, analysis of Cohen’s *d* effect sizes across patients revealed higher feature values at the peaks of dIEA cycles (**Figure 3A**). We observed significantly positive effect sizes for theta (sign test U(19) = 16, p = 0.004) and beta connectivity (U(19) = 17, p = 0.0007), as well as theta (U(19) = 16, p = 0.004) and alpha (U(19) = 16, p = 0.004) band power. Interestingly, we found a bimodal distribution of gamma band power effect sizes, indicating distinct patient subpopulations with higher gamma band power either at the peaks or troughs (**Figure 3B,S6**) Repeating the above analysis for RvF phases revealed that feature values were not preferentially higher in either phase, and the difference between rising and falling phases was not significant across the population in any feature (**Figure 3A, S7**). Our analysis of univariate features revealed no one electrophysiological biomarker that could strongly discriminate between the cycle peaks and troughs, but rather that subsets of features which discriminate between cycle phases are patient-specific.

**Figure 3.**
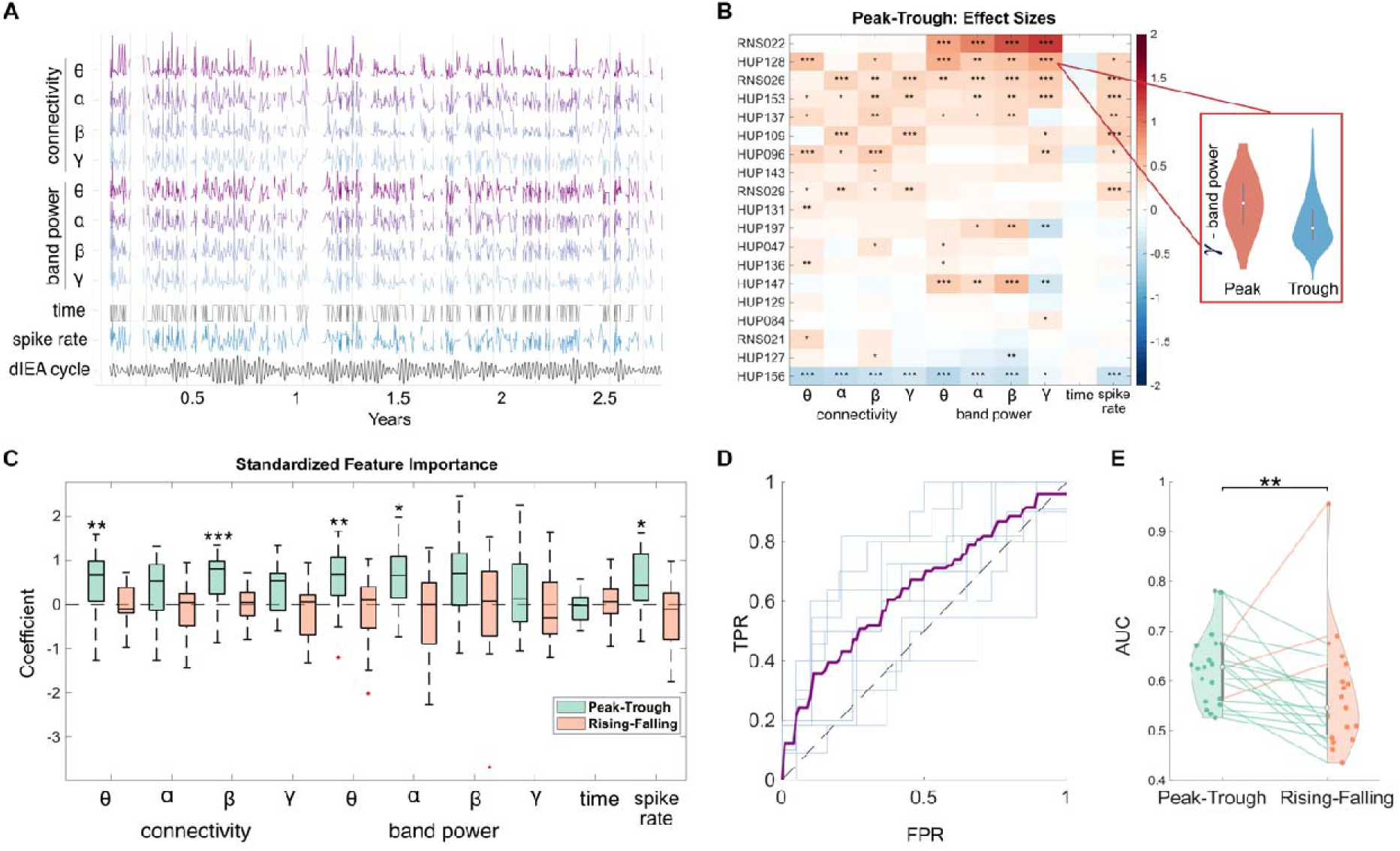
Association of background features to dIEA cycle phase. A) Evolution of feature values over time for an example patient, trend lines are omitted for ≥5 days without recorded SE data (HUP096). Features were smoothed for visualization purposes and normalized between clinical visit (vertical lines). B) Heatmap demonstrating population trends in PvT effect sizes. The relationship between electrophysiology and dIEA is an individualized phenomenon, with different patients having different features that best associate with dIEA cycle phase. C) Population distributions of PvT and RvF multivariate model coefficients. PvT models reveal increased activity and connectivity during the peak compared to the troughs of multidien dIEA cycles. **D)** Mean out-of-sample PvT prediction ROC curve for HUP096 (purple) and ROC for each fold (light blue). **E)** Population distributions of model performance (AUROC) between PvT and RvF models without spike rate. Model performance is significantly higher in PvT models.

### ML Model with background features

For each patient, we built binary linear SVM classifiers to predict cycle phase - PvT, and RvF - from the EEG features. Our models were able to predict PvT cycle phase above chance in all patients (AUC >0.5, 14/19 significant - **Figure S8, Table S1**), and RvF phase in 14/19 patients (8/19 significant - **Table S1**). Between the two classifiers, we saw higher performance in the PvT over the RvF model (sign test, p = 0.0044) (**Figure 3E**). Despite the fact that seizures preferentially occur in the rising phase of the cycle[5], our results indicate that EEG features are more strongly associated with the amplitude of dIEA cycles than individual phase bins.

Even after accounting for correlated features (**Figure S9**), spike rate, and time of day, model coefficients showed higher band power and connectivity in the brain during the peaks of the multidien dIEA cycles (**Figure 3C**). While there was between-patient variation in the magnitude of the coefficients corresponding to each feature, we identified significant population trends of higher connectivity in the theta (sign test U(19) = 17, p = 0.0007) and beta (U(19) = 17, p = 0.0007) bands, and higher power in the theta (U(19) = 16, p = 0.0044) and alpha (U(19) = 17, p = 0.0007) bands at the peaks of the cycles. We also correlated model performance with both dIEA cycle length and LE phase-locking value, and found no significant trends. This suggests that the representation of dIEA cycles in interictal baseline electrophysiology is independent of cycle length as well as LE entrainment (**Figure S10**).

### Phase dependence of stimulation response features

We hypothesized that multidien fluctuations would be temporarily superseded by the brain’s stimulation response, making multidien cycles more difficult to detect. Univariate analysis on post-stimulation periods was consistent with our univariate analysis of SE clips without stimulation, showing that brain connectivity and band power activity are increased at the peaks vs. troughs after stimulation (**Figure S11**).

As with our background EEG analysis, we used an SVM model to explore the multivariate relationship between post-stimulation features and phase. The PvT detection model achieved above-chance performance in all patients (median AUC 0.58 [0.55, 0.64], **Figure 4B**) and the RvF model achieved above chance performance in 12 of 17 patients (median AUC 0.53 [0.50, 0.60], **Figure S12**), indicating that multidien phase remained detectable in the seconds following stimulation. However, compared to PvT detection in scheduled events without stimulation, post-stimulation detection performance was reduced, suggesting a relative suppression of the relationship between the EEG and dIEA cycles. Finally, the standardized feature importance for both PvT and RvF models did not exhibit a significant directional effect at the group level (**Figure 4A**). In general, both the direction and magnitude of feature importance is more heterogeneous across patients in the post-stimulation detection scenario, when compared with background EEG model coefficients.

**Figure 4:**
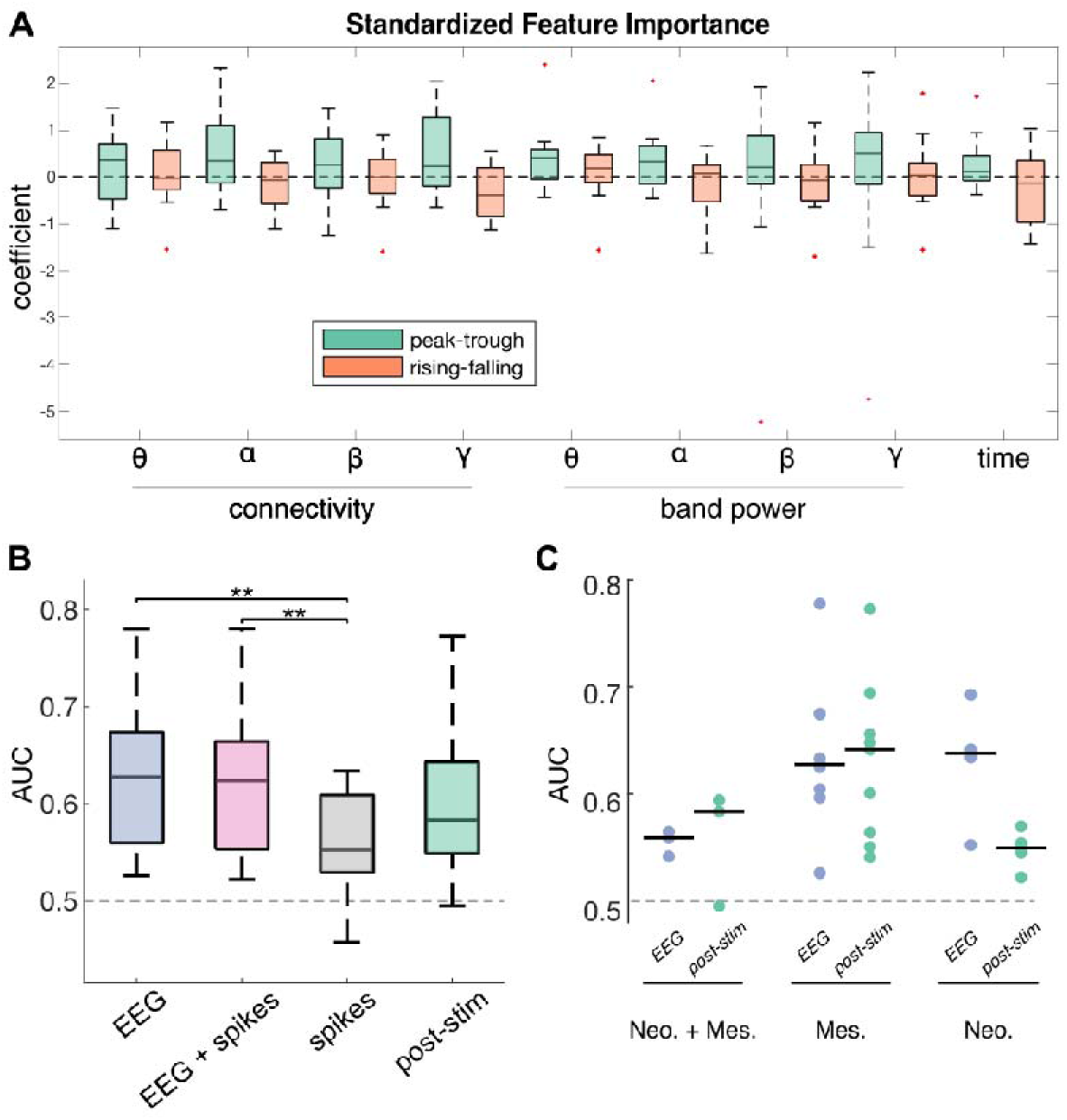
Post Stimulation Feature Importance and Model Comparison. **A)** Peak-Trough and Rising-Falling multivariate feature importance (Cohen’s D) in multivariate post-stimulation model. Stars indicate significance level of effect size distributions against a zero median (sign test). **B)** Comparison of PvT model performance (AUC) across SVM models trained with (1) background EEG features, (2) background EEG features + spikes, (3) spikes, (4) post-stimulation features. **C)** Comparison of EEG (no-stim) and post-stim model performance broken down by implant depth subgroups. Neo.: Neocortical implants, Mes.: Mesial temporal implants, Neo. + Mes.: one implant each in neocortical and mesial temporal structures.

### Comparison of ML model in different scenarios

Finally, we compared PvT performance for models trained with the following feature sets: (1) features from SE clips without stimulation, (2) features from SE clips without stimulation AND spike rate, (3) spike rate, and (4) features from post-stimulation windows. Across the four feature sets, we found an increased ability to detect peak vs. troughs in the EEG (0.63 [0.56 - 0.67]) and EEG + spikes (0.62 [0.55 - 0.66]) models when compared to the spike only (0.55 [0.53-0.61], rank sum p = 0.0038) and post-stimulation models (0.58 [0.55-.64], rank sum p = 0.205) (**Figure 4B**). We further analyzed model performance by implant depth, the decrease in performance between background EEG to post-stim models appeared to be driven by patients with neocortical implants (median AUC 0.64 - 0.55) (**Figure 4C**). Similarly, the superior performance of PvT models when compared with RvF models persisted in the EEG feature sets but not in the post-stimulation or spike rate only feature set (sign test p= 0.03, 0.03, 1, 0.47).

Although spike rate alone did not significantly indicate phase for all patients, we incorporated spike rate into the linear SVM of background features to determine whether a more complex interaction could better be associated with phase position. We found that the inclusion of spike rate did not substantially change the median AUC, though both the EEG and EEG with spikes models performed better than the spike rate only model, which had the worst performance across all models compared, indicating that the inclusion of background EEG features adds complementary information to phase detection (**Figure 4B**).

## Discussion

Our study investigates RNS EEG recordings in 20 patients to build on prior work examining the relationship between EEG features and multidien cycles of cortical excitability. We leverage the unique recording and stimulation characteristics of the RNS dataset to complete the largest investigation to date on neuromodulation and dIEA cycles. Specifically, we show that: (1) beyond spike rate, features of interictal EEG signals contain information pertinent to detecting dIEA phase, (2) EEG features that are most associated with cycle phase are patient-specific, and (3) after neurostimulation, the EEG-dIEA relationship is suppressed. These findings may have important implications for tailoring pharmacological and neurostimulation therapy to patient chronotype, as well as for using multidien cycles as a predictive biomarker of seizure risk and understanding their underlying mechanisms.

### Spike rate does not tell the whole story: even after accounting for spikes, broader EEG features are associated with cycles

Prior work investigating the multidien nature of epilepsy characterized the presence of cyclical seizure risk in acute event counts both in RNS dIEA[5], [12] as well as clinical spikes in different implantable devices[8]. We found evidence of multidien dIEA cycles in background EEG features beyond epileptiform spikes and acute signatures of epileptiform activity, and showed that multivariate EEG models outperformed spikes alone in predicting dIEA cycle phase. Our results suggest that there is complementary information between spikes and EEG features, and indicate that multidien cycles can be identified even below the spike detection threshold. It may be that some other latent mechanism, linked to both epileptiform activity and subthreshold EEG features, is responsible for cycle generation. EEG features could present an alternative or addition to using spikes in future therapeutic algorithms that incorporate patient chronotypes to titrate pharmacological or neuromodulatory therapy.

### Patient-specific features underlie the population level dIEA-EEG association

The mechanism that causes variability in cycle period between patients is unknown and could be driving the observed differences in important feature sets between patients. It is also possible that differences in implant locations and electrode type could drive inter-patient variability in feature importance, given that measures of brain activity vary across brain structures [23]–[25]. Subgroup analysis revealed that the EEG-dIEA cycle coupling is strongest in neocortical patients. Prior work has shown that within patients dIEA cycles are stable between anatomical regions[5] but our finding suggests that there may be an anatomical basis for the presence of cycles in background recordings. Future work should rigorously investigate sub population effects on multidien cycles in background EEG features.

### The relationship between dIEA cycle and EEG features is attenuated after neurostimulation

There is room for improving responsive stimulation specificity, and a functional question that arises from our observation of multidien cycles in epilepsy is: can stimulation perturb these cycles? Recently, Gregg et al. provided evidence that the power of the dominant dIEA cycle could be modulated by thalamic deep brain stimulation [7], and a broad body of literature has shown that stimulation acutely suppresses EEG signal features[17], [26], [27]. Our results are consistent with the potential for neurostimulation to disrupt cycles – we find a general suppression of the relationships between the EEG and dIEA with lower model effect sizes and limited population trends in model coefficients after stimulation, particularly with neocortical implants. One possible theory is that stimulation causes a stereotyped brain activity reset[27], reducing feature variance across cycle phases and model performance. Another theory is that the brain state triggering the stimulation is inherently decoupled from the dIEA cycles.

### Sampling bias in RNS recording types reflects cycle phase

We found that cycle phase caused an imbalance in the availability of scheduled events both with and without stimulation, potentially biasing analyses towards recordings with different levels of cortical excitability. Given that values of EEG features fluctuate with cycle phase at the group level, sampling bias should be a practical consideration for studies separately analyzing RNS recordings with or without stimulation.

### Limitations

Our study has several limitations. The relatively small size of our patient cohort, and the variability within it, precluded us from performing robust statistical sub-analyses or further exploring mechanistic hypotheses. Further studies with larger sample sizes are needed to confirm and expand upon our findings, and our group is currently working on a federated analysis of RNS data from multiple institutions to accomplish this goal [28]. We were also limited by the highly irregular sampling of EEG recordings by the RNS device, which prevented us from extracting cycles from EEG features using the same statistical tools and methods we applied to dIEA counts. This under sampling of the intracranial EEG is a fundamental challenge presented by this device. Future work investigating the observed phenomena should leverage continuous recording paradigms. Additionally, we select the cycle of interest based on LE PLV. While RNS LEs may not always capture true seizure activity, they have been shown to be correlated with electrographic events[29]. Despite these limitations, our study provides insight into the physiological correlates and underpinnings of the measured dIEA cycles in patients with RNS.

#### Declaration of anonymous identifiers

Sample/patient IDs disclosed in this study (HUPXXX, RNSXXX) are not identifiable and cannot be linked to any protected health information by any individuals outside of our research group.

#### Declaration of generative AI and AI-assisted technologies in the writing process

During the preparation of this work the authors used ChatGPT to edit for conciseness. After using this tool/service, the authors reviewed and edited the content as needed and take full responsibility for the content of the publication.

## Data Availability

Data associated with the RNS device was obtained through a data use agreement between the University of Pennsylvania and NeuroPace, and cannot be publicly shared. However, code used to process RNS device data and perform our analysis can be found on GitHub at https://github.com/penn-cnt/RNS_processing_toolbox and https://github.com/wojemann/multidien-cycle-biomarkers

https://github.com/wojemann/multidien-cycle-biomarkers

## Abbreviations

RNS: responsive neurostimulation
dIEA: device-detected interictal epileptiform activity
PLV: phase-locking value

## Acknowledgements

We thank Magda Wernovsky for her help in consenting RNS patients.

