## Supplemental Information for "Resting-state background features demonstrate multidien cycles in long-term EEG device recordings"

**Contents**

1. Supplemental Methods

2. Supplemental Figures

3. Supplemental Tables

**Supplemental Methods**

***RNS Device- Detection Criteria***

Onboard detectors track signal features (e.g. signal band power, line length, zero crossings) in up to two channels at a time, and are triggered when the feature exceeds a preset threshold over a specified time interval. Detectors are tuned by a licensed neurologist to be triggered by both short epileptiform discharges, if the detection threshold is exceeded briefly (usually < 2s), and by seizures, if the detection threshold is exceeded for a longer period of time (usually 10-30s).

**Patient Selection**

We excluded eight patients for the following reasons: insufficient IEA detection data (N=2), lack of a significant dominant multidien cycle (N = 3), insufficient scheduled EEG recording clips without stimulation (N=4), insufficient scheduled EEG recording clips with stimulation (N = 6, 3 patient exclusion overlap).

***dIEA Cycle Analysis***

We first removed the first 30 days of dIEA data to account for potential implant effects. We then normalized the hourly dIEA counts between neurologist visits to account for detection parameter changes (**Figure 1A**). Data gaps of 5 hours or less were linearly interpolated, otherwise data was segmented at larger recording gaps. We applied a Morlet wavelet decomposition to each continuously recorded segment to obtain a spectrogram (octaves: 12, minimum scale: 0.008). We averaged the spectrogram across time to obtain a time-averaged periodogram, then averaged periodograms across segments to obtain a single periodogram per patient (**Figure 1B**). We identified peaks in the time-averaged periodogram using MATLAB’s *findpeaks* function. Periods corresponding to peaks that rose above the 99% confidence interval of the Fourier red-noise null model were identified as significant dominant modes of the dIEA count signal (Karoly et al., 2020; Torrence & Compo, 1998).

For each dominant mode in the range of 3-60 days, we applied the inverse wavelet transform to reconstruct the component of the signal with the associated period (basis function: delta function, reconstruction factor: 0.776, decorrelation factor: 2.32, scale averaging factor: 0.60, energy de-scaling factor: -1/4) (Farge, 1992). Prior studies report that significant multidien cycles are not present in all subjects (Baud et al., 2018; Karoly et al., 2018) and five patients did not have a significant dominant mode within the 3-60 day window and were excluded from the study. For the remainder of our analysis, we selected the multidien period in each patient that corresponded most with seizure occurrence. That is, we used Long Episodes (LE) as a proxy for seizures and chose the multidien period with the highest phase-locking value of LE to the reconstructed (inverse wavelet) signal (Anderson et al., 2022; Baud et al., 2018) (**Figure 1B,C**). Finally, we used the Hilbert transform to obtain the instantaneous phase angle for each timepoint of the chosen reconstructed hourly dIEA cycle (**Figure 1C**).

***Spike Detection***

We built our spike detector with the following settings: data in each SE clip was filtered between 10 and 100 Hz using a 6th order butterworth bandpass filter. The channel-specific peak-to-peak spike amplitude threshold criteria was defined as the maximum of 7.5 standard deviations above the mean across all SE clips or 6.5 times the absolute median of the data within the clip. Setting dual thresholds ensured that the detector was not overly sensitive when clip amplitude was elevated. Additionally, the maximum and minimum spike peaks were required to fall within a window of 15 to 200 ms (**Figure 2A**). Any spikes that occurred within 1 second of a stimulation interval were excluded due to stimulation artifacts.

***Feature Calculation***

To calculate band-limited bandpower in each frequency band of interest (theta, alpha, beta, gamma) we applied the MATLAB *bandpower* function to the entirety of the recording window - either an entire scheduled event without stimulations or the window after stimulation. We then took the maximum band power value between the two detection channels in each frequency band to get one band power feature value per recording, per frequency. We excluded the delta frequency band due to onboard filtering settings in the RNS device that prevent analysis of this activity.

To calculate band-limited connectivity we first split the recording of interest into non-overlapping one second clips. We then, in each clip, calculated the cross-wavelet transform connectivity using the MATLAB function *xwt* (Torrence & Compo, 1998) between the two recording channels on the same electrode in each frequency band of interest providing one connectivity value per clip, per electrode, per frequency band. We then averaged across all of the clips to obtain an average connectivity value for each electrode in each frequency band and, in each frequency band, took the maximum value between the two electrodes. This provided one connectivity value per frequency band in each recording.

An important note in interpreting our EEG-dIEA results is that, while dIEA detection criteria can include thresholds of bandpower ranges as well as some spikes, our analysis investigates background signals explicitly when these detection criteria were not met - windows after detections or recordings without any detections.

***Machine learning modeling***

During univariate feature analysis, we observed a high correlation between feature sets (**Figure S9**), and thus applied a principal component analysis (PCA) transformation to the feature data in each fold to mitigate the multicollinearity (Massy, 1965). Only the first n principal components that explained 95% of the variance became our training features, in order to limit the destabilizing effect of principal components with smaller associated eigenvalues. We weighted the cost of misclassification by initializing our models using the class imbalance of the training fold, to prevent training-class imbalance from affecting our model generalizability. All predictions that we generated were out-of-sample predictions on the validation dataset for each fold, meaning that their accuracy and effect size is a better representation of how accurately the model is learning the relationship between the features and response variable.

To interpret the relationship between the features and response variable, we deployed an extension of principal regression analysis and applied the inverse PCA transform to the standardized model coefficients - scaled by the variance of the explanatory and response variables in each fold - to transform them back into feature space and compare across patients (Massy, 1965). Because we use a 5-fold cross-validation paradigm to assess the performance of our models, to get one feature importance value per patient-specific model we take the average feature value across k-folds. Analysis of the patient-specific model coefficients (**Figure S8**, **Table S1**) reveals that coefficients are highly consistent across k-folds. When we plot the distribution of model coefficients for both the stimulation-free and post-stimulation recordings we only plot the coefficients of the patients who had a validation AUC above 0.5. The reason for this exclusion is to prevent any coefficient inflation due to overfitting from biasing the population coefficient distributions.

**Supplemental Figures**

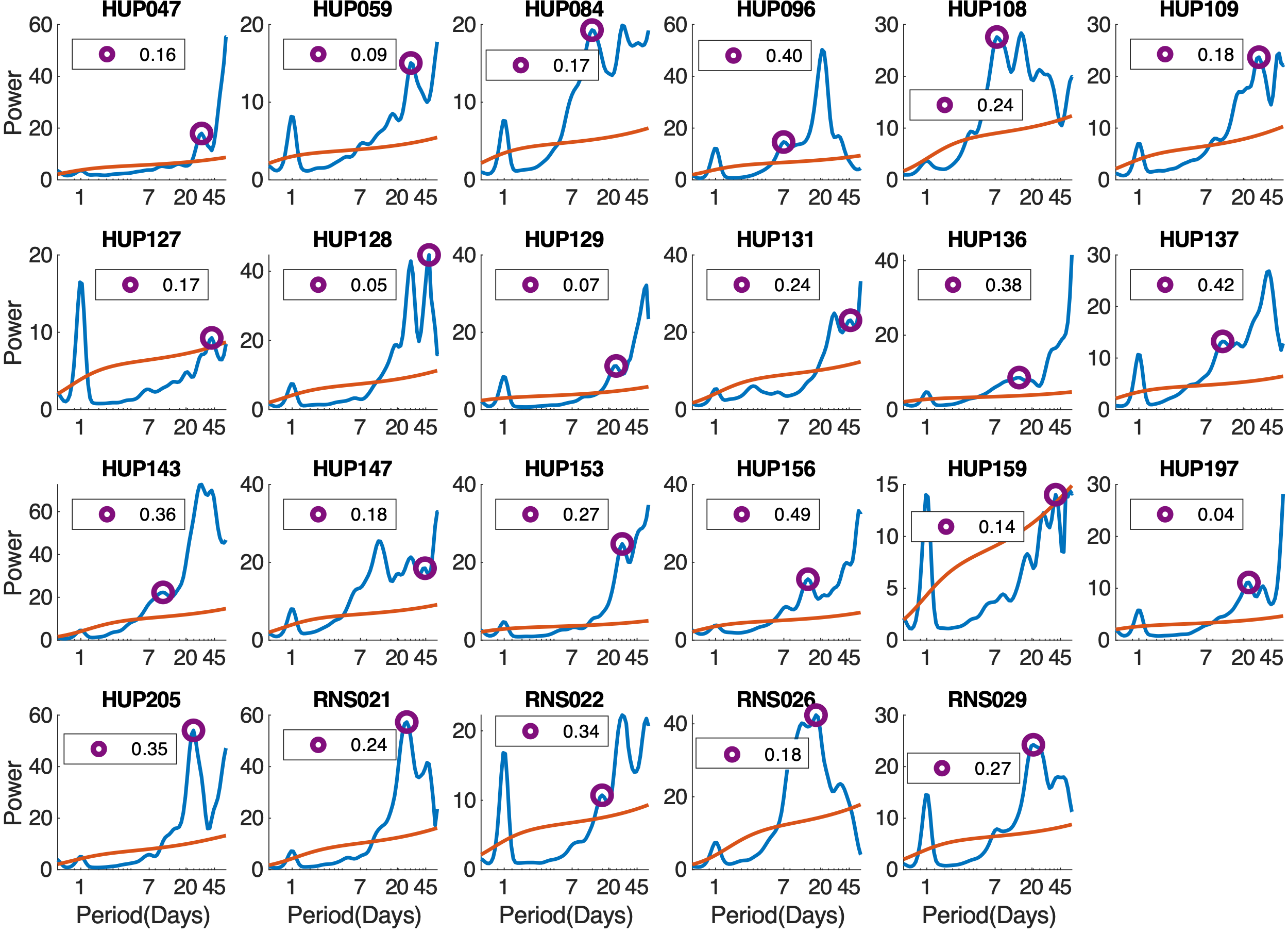

**Figure S1. Individual dIEA count Periodograms**. Periodograms showing the average power over time for a range of dIEA cycle periodicities. The red line denotes the 99% confidence interval of the red noise null model described in *Torrence and Compo et. al., 1998*. We fit the null model using first order autoregressive coefficients of the longest continuous segment of dIEA detections after normalizing between clinician visits. The purple circle marks the multidien peak that has the highest phase entrainment of Long Episodes among all multidien peaks, with the actual phase locking value of Long Episodes to cycle phase in the legend. Plots show inter-patient variability in terms of the prominence of circadian cycles as well as the period length of dIEA count dominant modes.

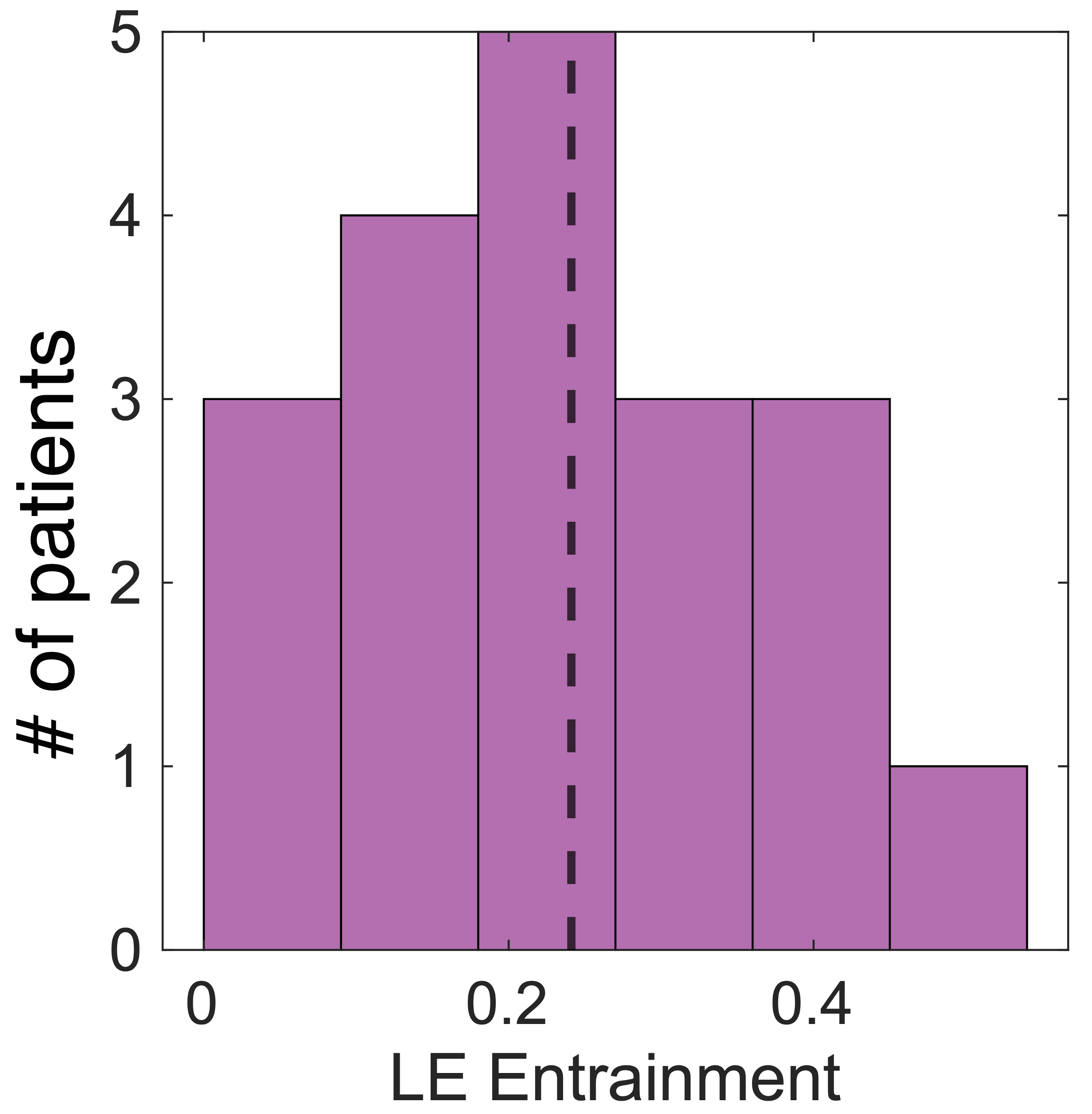

**Figure S2. Distribution of phase-locking values between long episode and multidien phase.** Distribution across all patients of LE phase locking value. Each sample in the distribution is the highest LE PLV across all dominant modes of the dIEA signal for each patient. LE PLV was calculated by taking the axial mean of the LE distribution across cycle phase (**Figure 1C**) using the circ-stat toolbox (Berens, 2009).

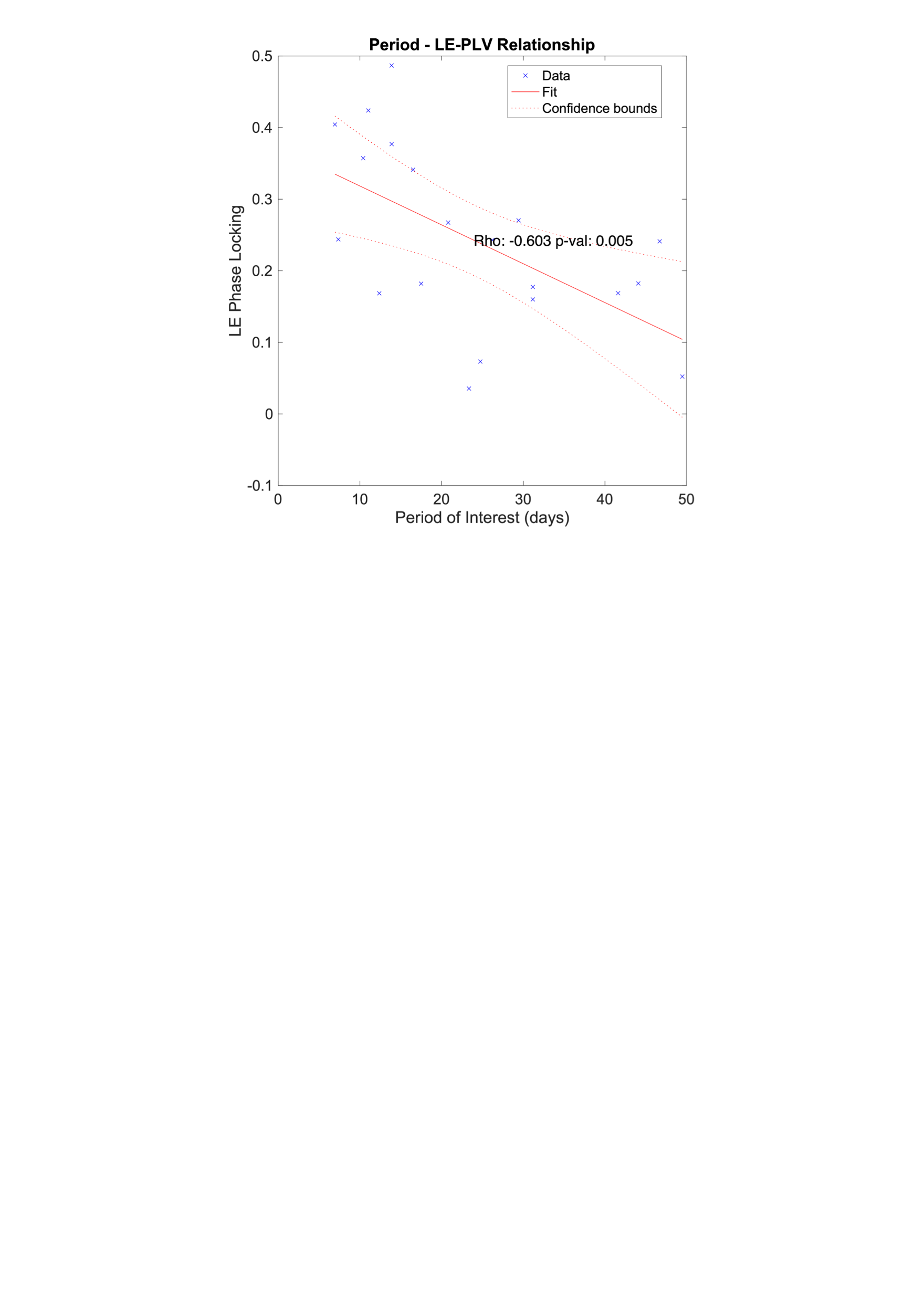

**Figure S3. Period-LE correlation.** Spearman rank correlation between the magnitude of LE phase locking and the period of the associated cycle. We analyzed the correlation between these two variables only for the highest LE phase locked cycle in each patient (**Figure S1**).

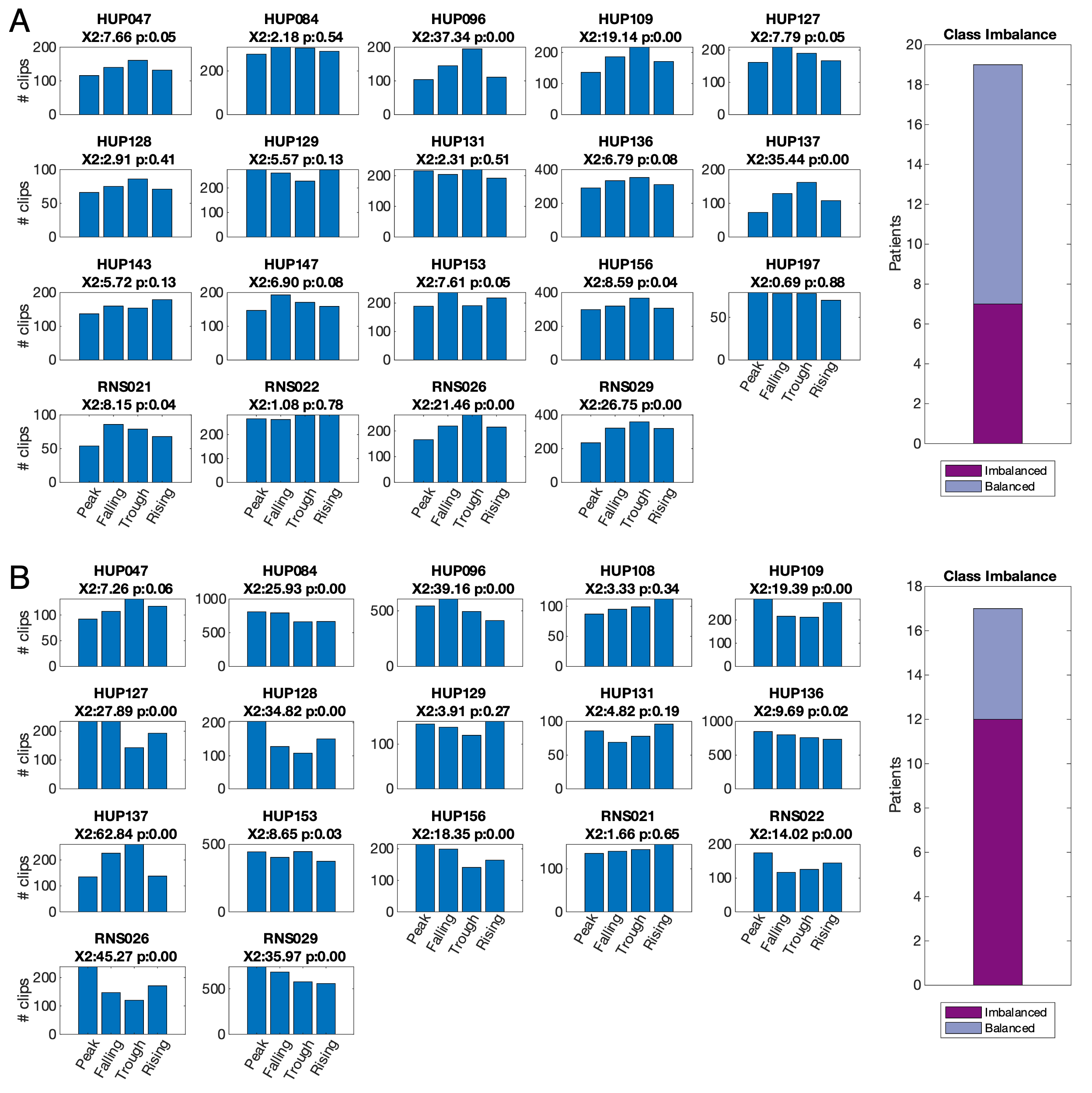

**Figure S4. patient-specific sample distributions. (A) scheduled events without stimulation, (B) scheduled events with stimulation.** Each subplot represents the distribution of recordings with (B) and without (A) stimulation across each of the four cycle phase categories. The bar plot to the right of each population plot shows the number of patients with a significantly imbalanced class distribution (p < 0.05) assessed by a chi-squared goodness of fit test to a uniform distribution. At the individual level, 7 of 19 patients demonstrated a significant imbalance in non-stimulation SE sampling and in 12 of 17 in post-stimulation SE sampling. Chi-squared test statistic and p value up to two decimal points shown in figure title. p: 0.00 is equivalent to p < 0.01.

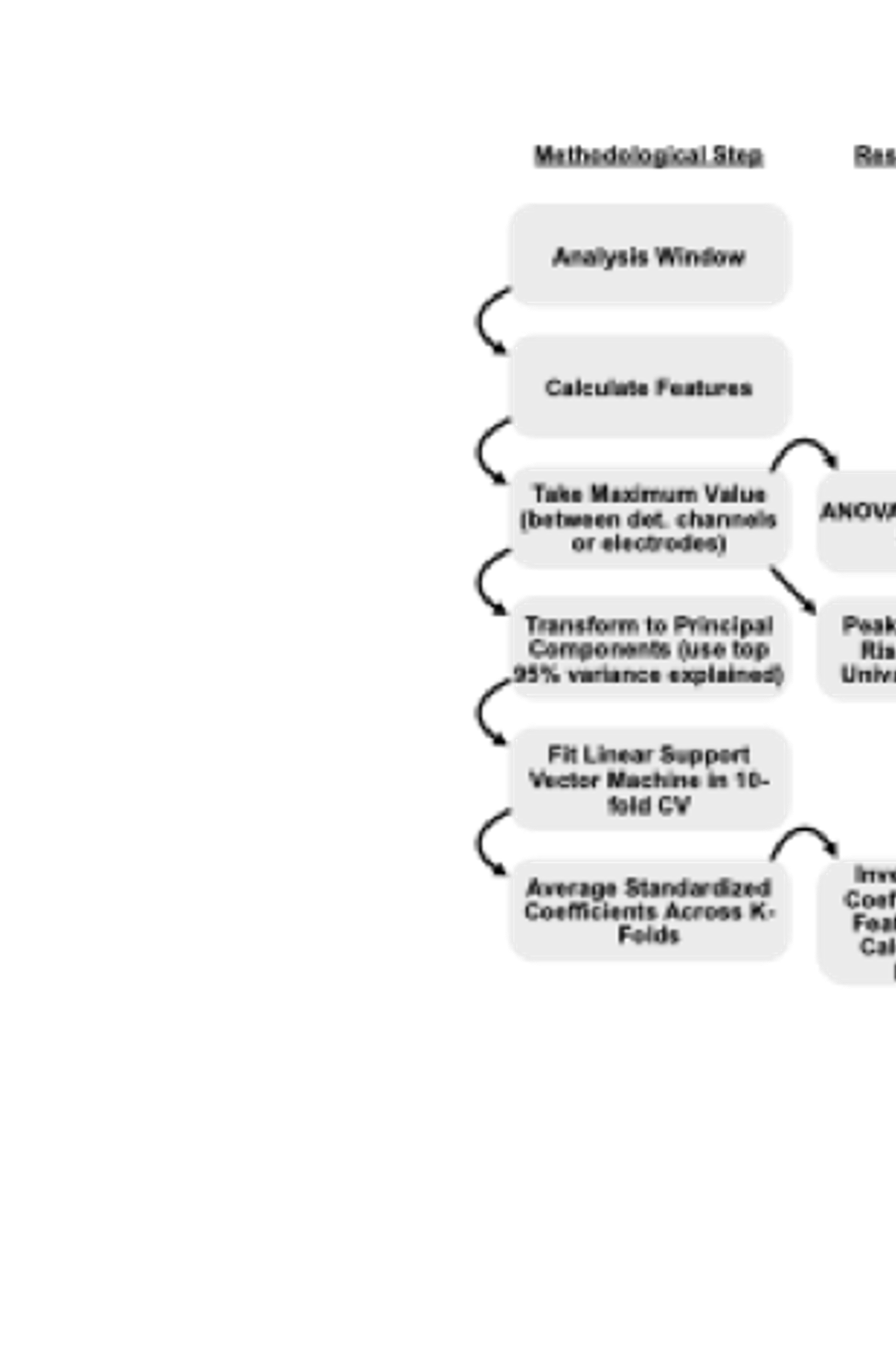

**Figure S5. Analysis Flowchart.** Analysis pipeline for both univariate and multivariate analyses in both non-stimulation SEs and post-stimulation windows. Flowchart shows the order of steps for calculating feature importance in two manners: univariate effect size, and standardized model coefficient. For multivariate analysis, we use the minimum required number of components to explain 95% of the variance in the data to remove principal components associated with spurious correlation between the features (Massy, 1965).

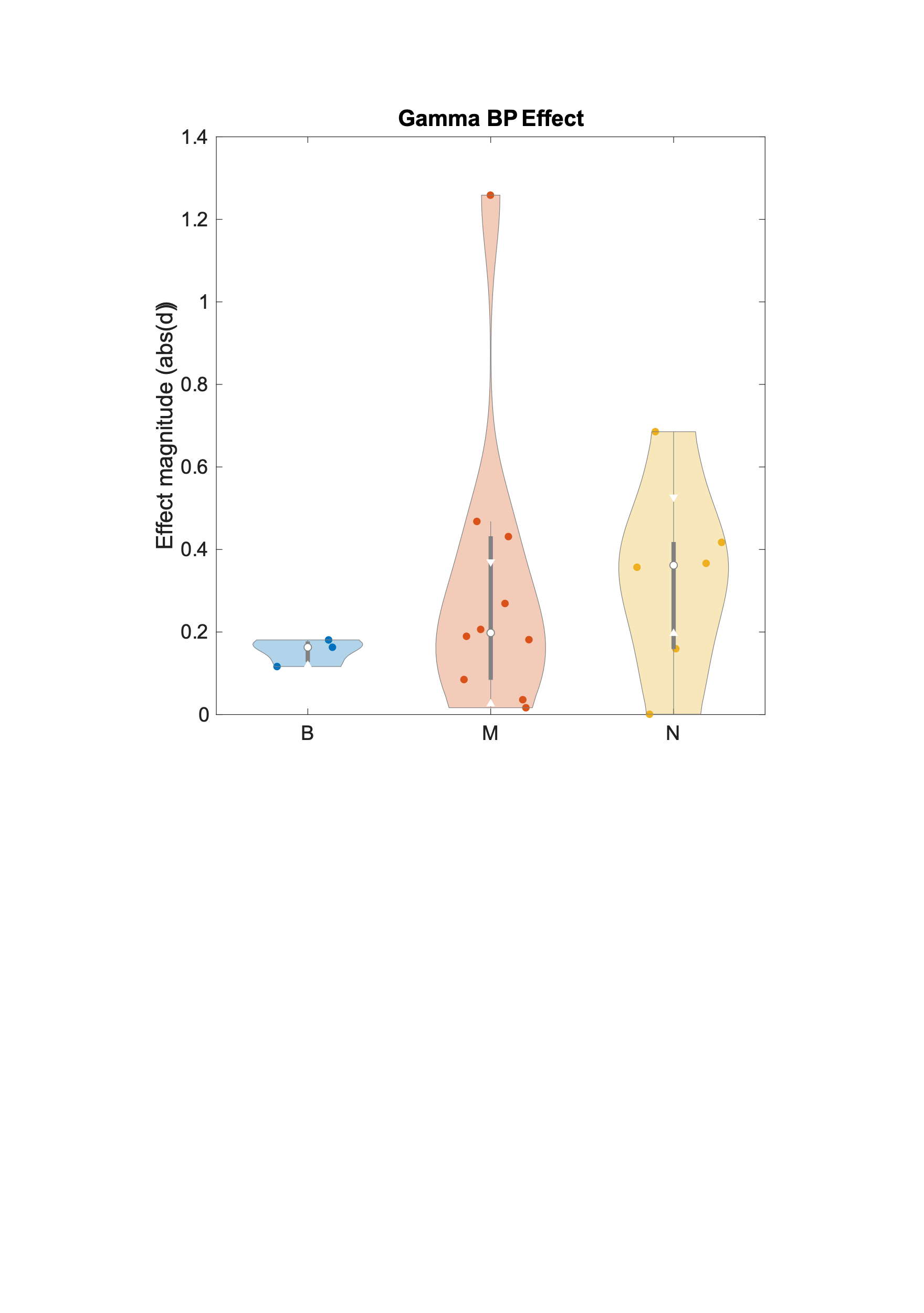

**Figure S6. Gamma effect size sub-groups.** Magnitude of gamma band power effect size grouped by patients with neo-cortical foci (N), mesial temporal foci (M) or both (B). When performing a sub-group analysis of the gamma-band power distribution of effect sizes, we observe a higher univariate feature importance (absolute effect size) in neocortical patients, but lacked the statistical power to rigorously examine this phenomenon

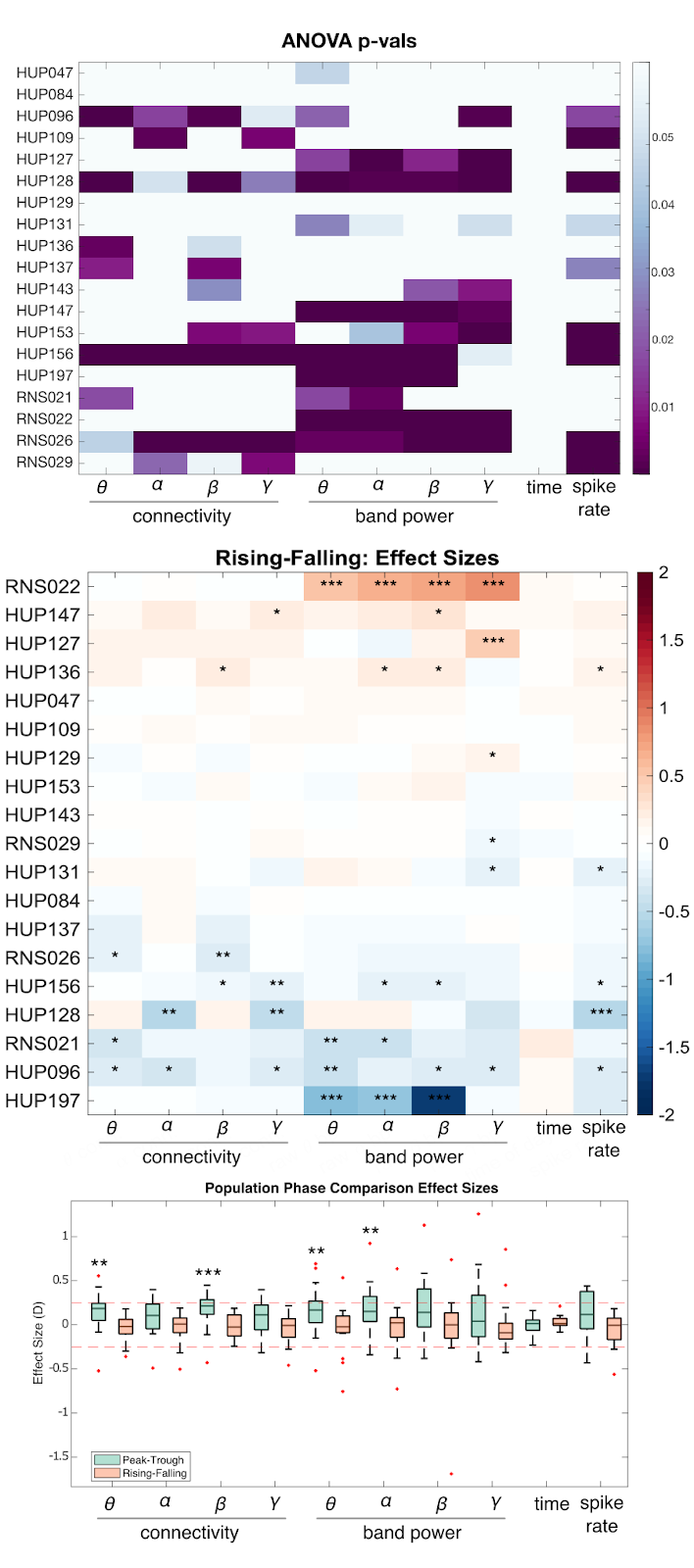

**Figure S7. Additional univariate analyses.** A) In each patient and for each feature, we used a one-way ANOVA to test the effect of phase group on feature value in the SE clips without stimulation. We found that 17 out of 19 patients possessed at least one feature which significantly discriminated between phase groups, with the most common significant features being theta (11/19), alpha (10/19), and gamma-band power (10/19). However, there was not any one clear feature that was generally strongly associated with phase for all patients. Additionally, the proportion of day and night recordings was similar across all phase groups, even in the presence of device recording limitations.

B) Rising-Falling effect sizes. We also calculated the difference in each feature distribution between the rising and falling phases of the cycles (peak-trough shown in **Figure 3B**). Significance of the separation in features between phase groups is denoted in stars (* p < 0.05, ** p < 0.01, *** p < 0.005). C) We lastly show the population distribution of effect sizes for each feature. Red-dashed lines indicate lower bound for moderate effect size (± 0.25). Stars indicate significance level of effect size distributions against a zero median (signtest)

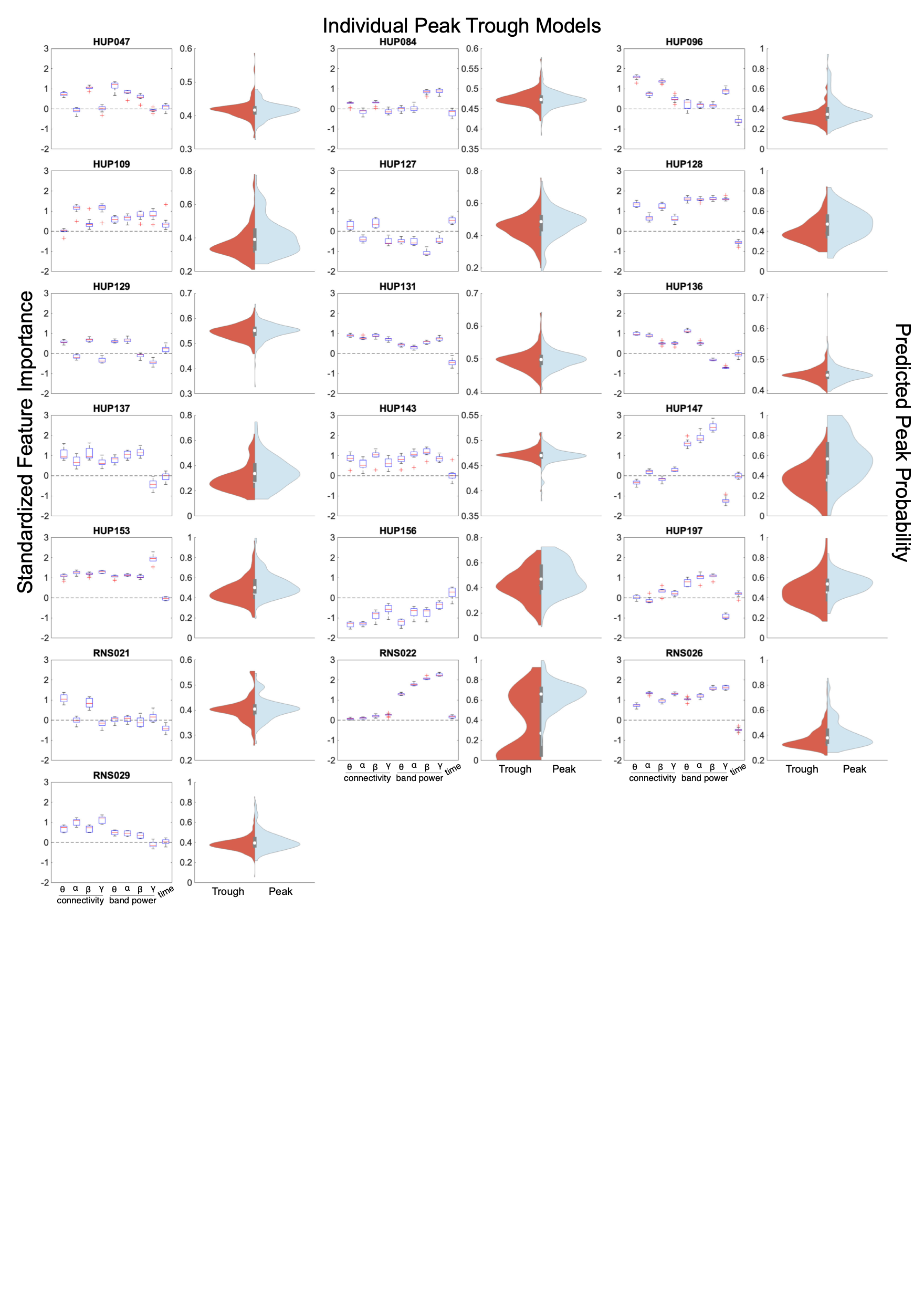

**Figure S8. Background EEG Model performances.** Patient level feature maps showing the distributions of linear SVM model coefficients across k-folds for peak-trough models. [0,1] scaled predicted probability distributions to the right of each feature distribution shows class separation with the true peak on the right and true trough samples on the left. The AUC and p-value associated with out of sample model performance for both the peak-trough (shown) and rising-falling models is shown in the patient information table.

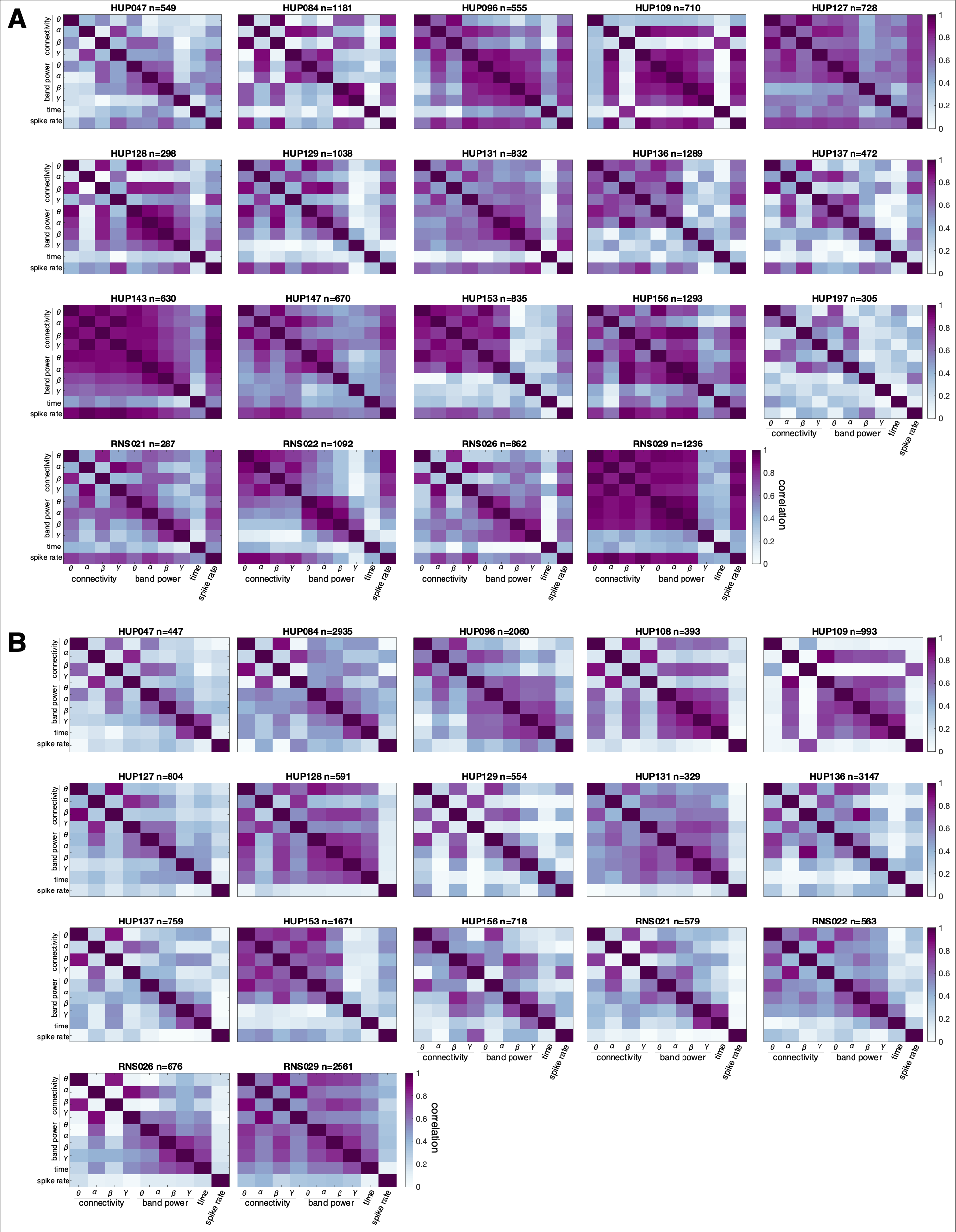

**Figure S9. Feature correlation.** Features from scheduled events without stimulation (A), and with stimulation (B). Qualitative group level analyses reveal that there is high multicollinearity between features in both the stimulation and no-stimulation groups, but which features are correlated is not necessarily consistent across patients.

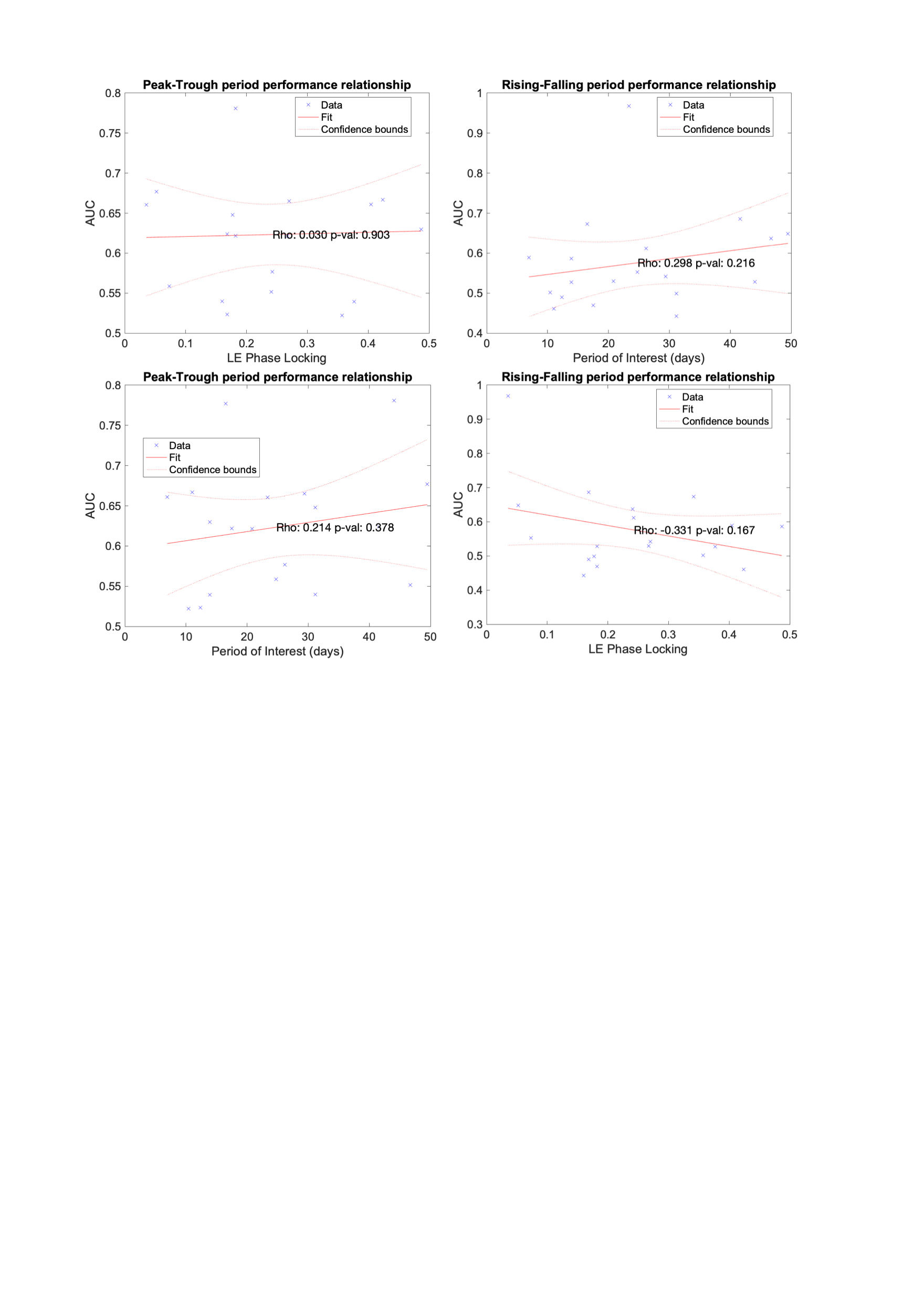

**Figure S10. Relationship between model performance and cycle period.** Despite the strong correlation between period of interest and LE phase locking, we observe no significant relationships between LE phase locking, length of cycle period, and model performance. Correlation coefficients are spearman correlation. The trend persists and all correlations are insignificant for the models with spikes as well.

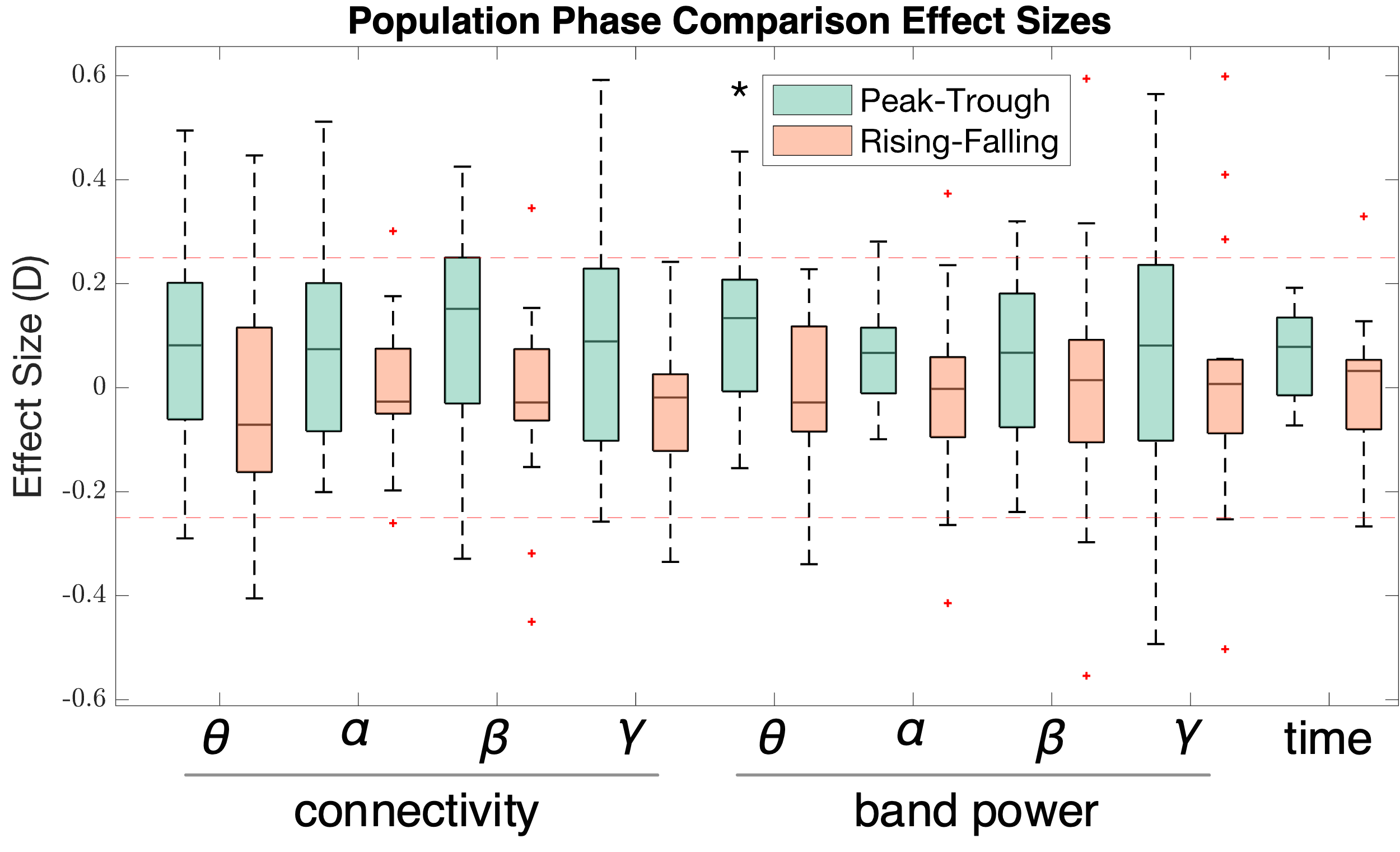

**Figure S11. Univariate Post-stimulation effect size distributions**

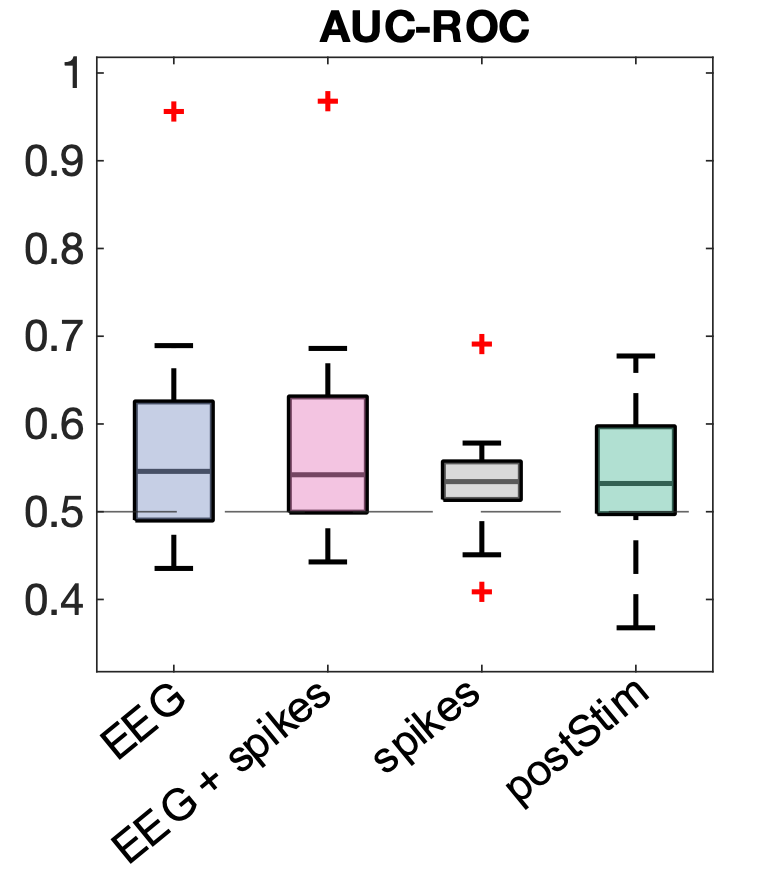

**Figure S12. ML model comparison for Rising-Falling Phases**

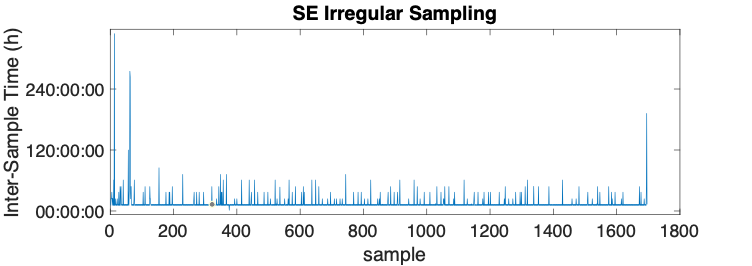

**Figure S13. Non-uniform sampling of SE recordings in HUP096**.

**Supplemental Tables**

**Table S1.  Patient-level multivariate model performance**

| **ID** | **PvT AUC** | **PvT p-val** | **RvF AUC** | **RvF p-val** |
| --- | --- | --- | --- | --- |
| HUP047 | 0.55 | 7.12E-01 | 0.44 | 4.52E-01 |
| HUP084 | 0.53 | 3.34E-01 | 0.49 | 5.63E-01 |
| HUP096 | 0.64 | **1.81E-04** | 0.59 | 9.95E-02 |
| HUP109 | 0.63 | **9.43E-05** | 0.48 | 4.50E-01 |
| HUP127 | 0.62 | **1.93E-04** | 0.69 | **9.72E-10** |
| HUP128 | 0.69 | **6.66E-04** | 0.65 | **2.29E-02** |
| HUP129 | 0.56 | **3.87E-02** | 0.55 | 1.30E-01 |
| HUP131 | 0.56 | 1.08E-01 | 0.63 | **9.25E-06** |
| HUP136 | 0.54 | 7.71E-01 | 0.53 | 5.47E-01 |
| HUP137 | 0.67 | **1.24E-04** | 0.48 | 2.65E-01 |
| HUP143 | 0.53 | 1.61E-01 | 0.51 | 3.97E-01 |
| HUP147 | 0.78 | **4.60E-16** | 0.51 | 7.25E-01 |
| HUP153 | 0.67 | **2.58E-08** | 0.57 | 1.08E-01 |
| HUP156 | 0.63 | **1.26E-08** | 0.59 | **4.06E-04** |
| HUP197 | 0.67 | **4.15E-04** | 0.96 | **4.39E-21** |
| RNS021 | 0.60 | 5.59E-01 | 0.60 | 3.78E-01 |
| RNS022 | 0.78 | **2.42E-29** | 0.67 | **3.29E-12** |
| RNS026 | 0.63 | **1.84E-05** | 0.46 | 4.55E-01 |
| RNS029 | 0.60 | **1.42E-04** | 0.53 | **1.29E-02** |
